# sEEGnal: an automated EEG preprocessing pipeline evaluated against expert-driven preprocessing

**DOI:** 10.64898/2026.04.16.26351021

**Authors:** Federico Ramírez-Toraño, Christoffer Hatlestad-Hall, Ainar Drews, Hanna Renvall, Paolo María Rossini, Camillo Marra, Ira H. Haraldsen, Fernando Maestu, Ricardo Bruña

**Affiliations:** Center for Cognitive and Computational Neuroscience, Universidad Complutense de Madrid, Madrid, Spain; Department of Neurology, Oslo University Hospital, Oslo, Norway; University of Oslo, UiO: Management and Support Units, IT Department; BioMag Laboratory, HUS Medical Imaging Centre, Helsinki University Hospital, Helsinki University and Aalto University School of Science, Helsinki, Finland; Department of Neuroscience and Biomedical Engineering, Aalto University, Helsinki, Finland; Department of Neuroscience and Neurorehabilitation, IRCCS San Raffaele, Rome, Italy; Memory Clinic, Fondazione Policlinico Universitario Agostino Gemelli IRCCS, Rome, Italy; Department of Psychology, Catholic University of the Sacred Heart, Rome, Italy; Department of Experimental Psychology, Cognitive Psychology and Speech & Language Therapy, Complutense University of Madrid, Madrid, Spain; Health Research Institute of the Hospital Clínico San Carlos (IdISSC), Madrid, Spain; Department of Radiology, Complutense University of Madrid, Madrid, Spain

**Author notes:** Corresponding author Ricardo Bruña -Department of Radiology, Complutense University of Madrid, Madrid, Spain. These authors contributed equally to the work (Federico Ramírez-Toraño, Ricardo Bruña).

**Keywords:** EEG preprocessing, Automated pipeline, BIDS, Bad channel detection, Artefact detection

## Abstract

Electroencephalography (EEG) preprocessing is a critical yet time-consuming step that often relies on expert-driven, semi-automatic pipelines, limiting scalability and reproducibility across large datasets. In this work, we present sEEGnal, a fully automated and modular pipeline for EEG preprocessing designed to produce outputs comparable to expert-driven preprocessing while ensuring consistency and computational efficiency. The pipeline integrates three main modules: data standardisation following the EEG extension of the Brain Imaging Data Structure (BIDS), bad channel detection, and artefact identification, combining physiologically grounded criteria with independent component analysis and ICLabel-based classification.

Performance was evaluated against manual preprocessing performed by EEG experts at two complementary levels: preprocessing metadata (bad channels, artefact duration, and rejected components) and EEG-derived measures. In addition, test–retest analyses were conducted to assess the stability of the pipeline across repeated recordings.

Results show that sEEGnal achieves performance comparable to expert-driven preprocessing while preserving key neurophysiological features. Furthermore, the pipeline demonstrates reduced variability and increased consistency compared to human experts. These findings support sEEGnal as a robust and scalable solution for automated EEG preprocessing in both research and large-scale applications.

## 1. Introduction

Electroencephalography (EEG) is a non-invasive neuroimaging technique that measures the spontaneous electrical activity of the brain using a set of electrodes placed on the scalp[1]. The standard EEG setup typically includes a cap with integrated electrodes, digital amplifiers, and software for data processing and visualization[2]. Compared to other noteworthy neuroimaging techniques, EEG presents some vital advantages: high temporal resolution; fair spatial resolution with the modern high-density EEG setups; portability; significantly lower hardware and consumables expenses; non-invasiveness; and no radiation[3]. As a result, modern EEG has become one of the most promising neuroimaging techniques for aiding in the diagnostics of neurological disorders, characterizing mood disorders and mental conditions, and revealing and defining the neurophysiological underpinnings of human cognitive functions.

Importantly, the raw EEG signal captures not only neuronal activity related to brain functions, but also physiological artefacts originating from blinks and eyes movements, heartbeats, and body movements. Additionally, it captures non-physiological artefacts induced by electronic noise and alternating current (AC) power lines[4]. A “pre-processing” pipeline is a set of methods applied to EEG recordings to remove (or at least minimise) the effects of such artefacts, ideally keeping the desired brain activity unaffected. The pre-processing pipeline is an indispensable step before analysing EEG recordings to obtain valid results. Although there are general guidelines to this aim[5,6], they must be adapted to suit each specific experiment or condition.

The most common approaches of EEG pre-processing involve semiautomatic pipelines supervised by EEG experts. These approaches assure the quality of the pre-processed EEG data but present some considerable challenges in large-scale data analysis settings: the process presents an unaffordable time-consuming nature; the outcomes are strongly dependent on the expert who has performed the analysis; and, due to the dependence of the solutions on programming frameworks (mainly MATLAB and Python), the approaches are only suitable for field experts with some level of programming skills. In response to these limitations, several automated pipelines have been proposed, typically prioritizing either standardised preprocessing workflows or data-driven artefact detection strategies. However, many of these approaches do not explicitly integrate both perspectives within a unified and interpretable framework.

Here we present sEEGnal, an automated toolbox defined in separate modules, namely standardisation, bad channel detection, and artefact identification. The standardisation module is based on the EEG extension of the BIDS framework[7] to ensure structured, interoperable, and reproducible data handling. The bad channel detection and artefact detection modules combine physiologically grounded criteria with ICA-based decomposition. This design enables a transparent and modular preprocessing workflow that aligns with expert practices while facilitating scalability across datasets. Importantly, we evaluate sEEGnal against manual preprocessing performed independently by multiple EEG experts, allowing performance to be interpreted in the context of expert-driven variability. Given the absence of ground truth in real-world EEG data, performance is assessed at two complementary levels: preprocessing metadata (e.g. bad channels, artefact duration, and rejected components) and EEG-derived measures. We assess the impact of preprocessing on downstream signal properties, including power spectral density and functional connectivity, as well as the stability of the pipeline across repeated recordings within the same session. Altogether, these analyses provide a structured and multi-level validation of the proposed approach.

## 2. sEEGnal overview

The main characteristics of sEEGnal can be summarized as follows:

- **Comprehensive**. We have defined the algorithm underneath sEEGnal following widely accepted guidelines for EEG experiments[5,6]. The workflows and parameters associated with each module have been designed to obtain the best performance regarding outcomes while keeping time and memory consumption in mind.
- **Automatic**. We have designed sEEGnal to be completely automated, using a pre-defined configuration. At any moment, the user may change at convenience the configuration parameters stored in the corresponding JSON files.
- **Comparable to human EEG experts**. We have designed sEEGnal to match the performance of a human EEG expert, which is widely recognized as the current gold standard for EEG pre-processing.
- **Time and memory efficient**. We have developed sEEGnal to work efficiently on large datasets. To this end, in addition to its automatic nature, the efficiency in the use of resources (i.e. efficiency in computational and memory use) is vital.
- **Open-Source**. We have designed sEEGnal as an open-source tool, enabling free and unrestricted use within the community. The majority of the codebase is implemented in Python, while two modules are written in C to ensure adequate computational performance. We have developed sEEGnal in Python 3.12.10 and build upon the MNE-Python framework[8]. A complete list of dependencies is provided in the Supplementary Material.

The following subsections describe the high-level modules comprising sEEGnal: standardisation, bad channels identification, and artefact detection.

### 2.1. Standardisation

To facilitate the creation and distribution of public EEG datasets, and interoperability with concurrent EEG pre-processing and analysis frameworks, the first module focuses on the standardisation of the data following the EEG extension of the BIDS standard[7]. BIDS defines a consistent protocol for folder structure, filenames, metadata, and file formats to store several types of neuroimaging data (as well as other related information), including EEG, magnetoencephalography (MEG), and other neuroimaging data. An overview of the BIDS structure used in this pipeline is provided in the Supplementary Material.

### 2.2. Bad channel detection

Under our rationale, a bad channel refers to an electrode whose signal is unreliable due to technical issues rather than reflecting physiological brain activity. Neural signals tend to spread across electrodes because of volume conduction, resulting in spatially correlated patterns. In contrast, noise arising from electrode malfunction, poor contact, or hardware-related problems is often spatially isolated and does not propagate to neighbouring channels. Therefore, the identification of bad channels relies on detecting signals that deviate from the expected spatial and statistical structure of the EEG recording. Following this rationale, our pipeline defines bad channels as those exhibiting abnormal signal characteristics relative to the rest of the recording, including indicators of poor electrode contact, non-physiological amplitude, spectral abnormalities, artificial inter-channel coupling, or excessive variance:

- **High impedance**. The impedance between the skin and the electrode is a fast and accessible proxy for data quality, but only in conjunction with other metrics[9]. sEEGnal marks as bad those channels which impedance surpasses a predefined value. This value is configurable by the user through the JSON configuration files, with the default value set to 200 kΩ (see Supplementary Material).
- **Impossible amplitude**. The impossible amplitude refers to channels with low or high amplitudes that physiologically cannot reflect to brain signals. sEEGnal estimates the standard deviation of each channel and those channels with standard deviations outside a lower and upper thresholds are marked as bad. The lower and upper values are configurable by the user through the JSON configuration files, with the default values set to 1 µV and 500 µV.
- **Power spectrum**. An abnormal power spectrum between 45 and 55 Hz is a robust measure to detect possible bad channels (see Supplementary Material). First, sEEGnal estimates each channel’s power spectrum between 45 and 55 Hz using the Welch method. Then, for each channel, sEEGnal estimates the average power spectrum of the EEG recording excluding the current channel and compare the value of the current channel with the value of the rest of channels. sEEGnal marks as bad those channels with values 3 times greater than the average. This value is set by default but can be changed in the JSON configuration file.
- **Gel bridges**. A gel bridge occurs when conductive material (typically electrode gel) unintentionally connects two or more adjacent electrodes, creating an artificial and abnormal low-impedance pathway between them. This bridging causes the affected electrodes to record highly correlated or nearly identical signals[10]. sEEGnal estimates both the physical distance between each pair channels and the correlation coefficients of their data. All pairs of channels closer than 5 cm (the average distance between neighbouring sensors of the montages used for this study) that presents correlations above 0.999 are marked as bad channels. These values are set by default but can be changed in the JSON configuration file.
- **High amplitude variance**. Channels that present abnormal amplitude standard deviation when compared to the rest of the EEG channels are very likely bad channels. First, sEEGnal estimates each channel’s standard deviation. Then, for each channel, sEEGnal estimates the average standard deviation of the EEG recording excluding the current channel and compare the value of the current channel with the value of the rest of channels. sEEGnal marks as bad those channels with values 10 times greater than the average. This value is set by default but can be changed in the JSON configuration file.

The workflow of the module is presented Figure 1 and described as follow:

1. Load the raw EEG recording.
2. Apply a minimum pre-processing to the EEG recording to fulfil the ICLabel[11] requirements: define montage; a FIR bandpass filtering between 1 to 100 Hz; a downsampling to 500 Hz; a FIR notch filter at 50 Hz and harmonics; the segmentation into 4-seconds epochs; the application of an average re-referencing. No epochs were removed in this step.
3. Decompose the minimum-pre-processed EEG data into Independent Components (ICs) using a Second Order Blind Identification (SOBI) algorithm and classified them using the Python package MNE-ICALabel [12]. No explicit dimensionality reduction or rank correction was applied after re-referencing. Therefore, the effective rank of the data was implicitly determined by the re-referencing procedure.
4. Apply the bad channel criteria stepwise in the following order: high impedance, physiologically unattainable amplitude, power spectrum, gel bridges, and high variance. Channels marked as bad by either of these criteria are excluded from subsequent criterion evaluations.
5. Update the metadata accordingly to the results.

**Figure 1.**
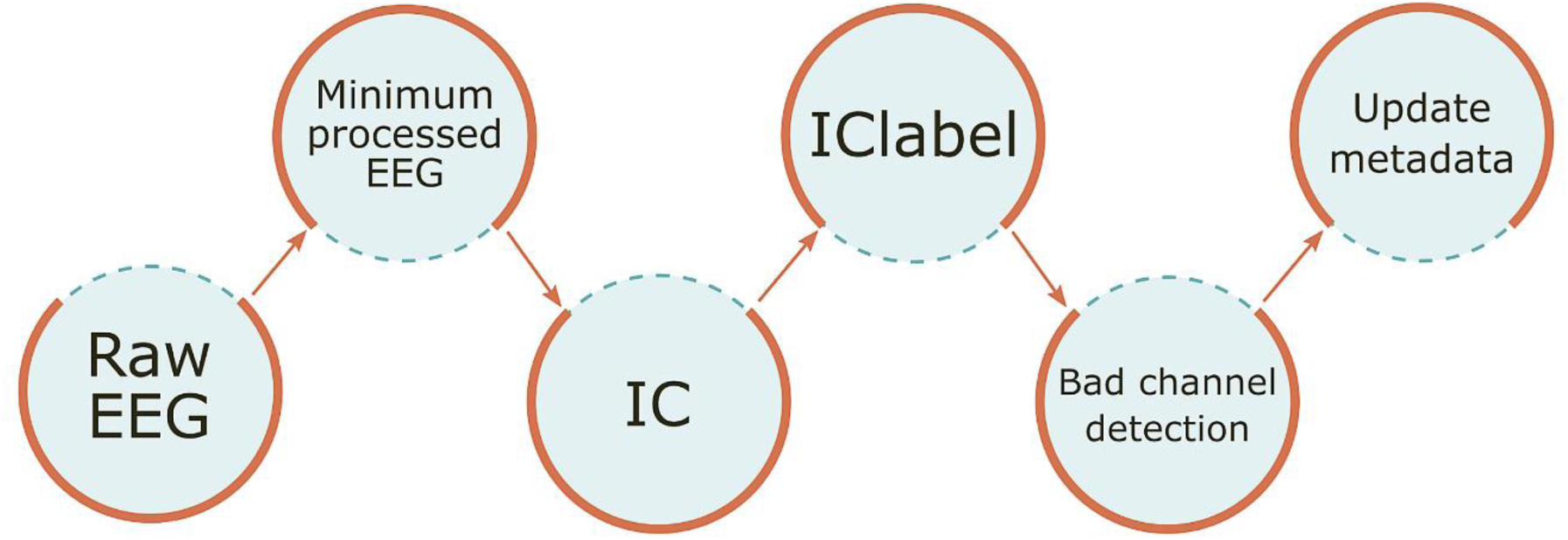
Sub-modules in Bad channel detection module. Abbreviations: IC, independent component.

### 2.3. Artefact detection

This module aims to identify artefacts present on EEG recordings, i.e. defining the onset and duration of the artefact. Specifically, eye-related, muscle-related, sensor-related, and others[13]:

- **Eye-related artefacts**. Eye-related artefacts arise primarily from electrooculographic (EOG) activity generated by eye blinks, vertical and horizontal eye movements, and eyelid flutter. These movements produce large-amplitude deflections that typically dominate the EEG below approximately 2–4 Hz, with maximal expression in frontal and frontopolar electrodes. To detect these artefacts, sEEGnal filters the EEG time series between 1 and 5 Hz and divided it into frontal channels and “background” channels. sEEGnal estimates the average standard deviation of both groups, and it looks for peaks in the frontal group with amplitudes at least 10 times greater than the “background” group. Finally, sEEGnal merges all the peaks to create the ocular artefacts. The threshold and the frequency limits are set by default but can be changed in the JSON configuration file.
- **Muscle-related artefacts**. Muscle artefacts arise from the contraction and mechanical tension of muscles located in proximity to the recording electrodes, producing electromyographic (EMG) activity that contaminates the EEG signal. These artefacts exhibit a broad spectral distribution, extending from near 0 Hz to frequencies exceeding 200 Hz, and their amplitude and waveform morphology are strongly influenced by the intensity, extent, and duration of the underlying muscle activity. To detect these artefacts, sEEGnal reconstructs time series using the ICs marked as “muscle” by ICLabel with a certainty threshold of 0.7. Then, sEEGnal filters the muscle time series for high frequencies between 110 to 145 Hz (a range commonly used as a heuristic for EMG artefact detection[8,14]), and, for each reconstructed channel, sEEGnal looks for peaks whose amplitude were 10 times greater than its average amplitude. Finally, sEEGnal merges all the peaks temporally closer than 0.5 seconds to create the muscle artefacts. The frequency limits, the certainty threshold and the distance between peaks are set by default but can be changed in the JSON configuration file.
- **Sensor-related artefacts**. Sensor-related artefacts—commonly termed channel jumps, electrode pops, or impedance shifts—originate from abrupt changes in electrode–skin impedance or mechanical instability of the sensor or cable. These artefacts manifest as sudden, large-amplitude steps or transient bursts, often with no physiological correlates. Their spectral signature is broadband but irregular, frequently introducing atypical high-amplitude low-frequency energy (< 5 Hz) due to the step-like displacement in the time domain. To detect these artefacts, sEEGnal filters the EEG time series between 0.5 and 5.0 Hz. Then, for each channel, sEEGnal estimates the standard deviation of the channel, and sEEGnal looks for peaks with amplitudes that are at least 5 times greater than the standard deviation of the channel. The threshold and the frequency limits are set by default but can be changed in the JSON configuration file.
- **Other artefacts**. This category encompasses signal segments with amplitudes or morphologies that are incompatible with known neurophysiological processes yet cannot be confidently attributed to a specific artefact source. These episodes typically present as abrupt, high-amplitude transients or sustained deviations that exceed biologically plausible voltage ranges for scalp EEG. Although their characteristics suggest non-neural contamination—such as sudden impedance fluctuations, cable disturbances, or undocumented environmental interference—the precise origin remains indeterminate. To detect these artefacts, sEEGnal filters the EEG time series between 2 and 45 Hz. For each channel, sEEGnal looks for time points with impossible high amplitudes, i.e. time points where the amplitude was above 500 µV. The threshold and the frequency limits are set by default but can be changed in the JSON configuration file.

The workflow of the module is presented in Figure 2 and described as follow:

1. The raw EEG recording is loaded.
2. Apply a minimum pre-processing to the EEG recording to fulfil the ICLabel[11] requirements: define montage; a FIR bandpass filtering between 1 to 100 Hz; a downsampling to 500 Hz; a FIR notch filter at 50 Hz and harmonics; the segmentation into 4-seconds epochs; the exclusion of channels marked as bad by the Bad channel detection module; the application of an average re-referencing. No epochs were removed in this step.
3. Decompose the minimum-pre-processed EEG data into Independent Components (ICs) using a SOBI algorithm and classified them using ICLabel using the Python package MNE-ICALabel [12]. No explicit dimensionality reduction or rank correction was applied after re-referencing. Therefore, the effective rank of the data was implicitly determined by the re-referencing procedure and the exclusion of bad channels.
4. Apply the artefact criteria stepwise for the following categories: muscle-related, sensor-related, and other.
5. Since muscle and sensor artefacts may dominate on the estimation of ICs, we perform a second IC estimations discarding epochs with artefacts.
6. Apply again the artefact criteria stepwise for the following categories: muscle-related, sensor-related, other, and eye-related.
7. Update the metadata accordingly to the results.

**Figure 2.**
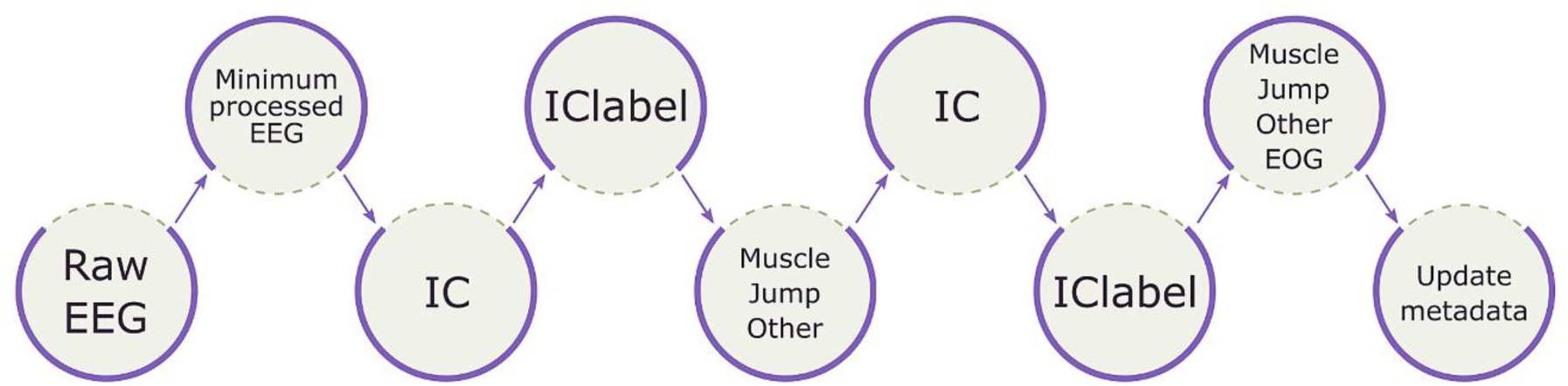
Sub-modules in Artefact detection module. Abbreviations: IC, independent component.

## 3. Material and Methods

Since there is neither an established gold standard for automated EEG pre-processing, nor a ground-truth associated to a real-world EEG recording, the quality assessment of an EEG pre-processing pipeline is not straightforward. To date, the best approach to evaluate the performance of an automatic EEG pre-processing pipeline is to consider EEG experts the gold standard and then compare the outcomes of the automatic tool with them. An overview of the process followed in this study to achieve our goal is presented in Figure 3.

**Figure 3.**
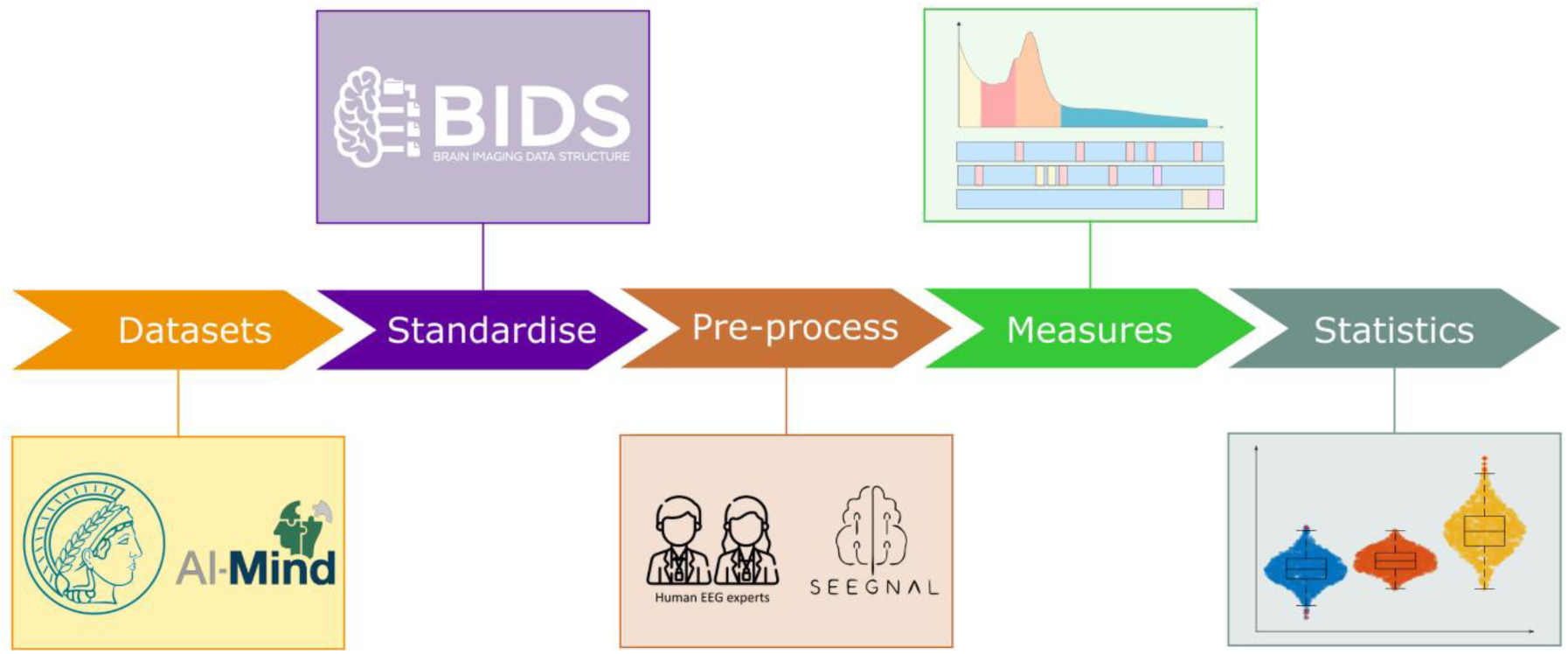
Overview of the pipeline followed in this study.

To use the EEG experts as gold standard in the comparison, we selected EEG datasets that fulfil two vital requirements: the recordings come from humans (not synthetic data), and the recordings have been manually pre-processed by EEG experts (or at least, visually inspected by them). In addition, we defined a comprehensive battery of outcomes to compare the performance of sEEGnal with the pre-processing by EEG experts.

We divided these outcomes into two conceptual categories, namely pre-processing metadata and EEG-derived measures. The pre-processing metadata refers to objective metrics of the EEG pre-processing, namely time and memory consumption, as well as the average number of bad channels, artefacts, and components rejected per EEG recording. As EEG-derived measures, we have selected two widely used EEG features, namely power spectrum, and functional connectivity.

The design of the pipeline and the development of the first versions started in 2022 as part of the European project AI-Mind [15]. The pre-processing of the datasets by sEEGnal was performed during 2024. The pre-processing of the dataset by the EEG experts was performed during 2025. The pre-processing of both datasets and the performance assessment was done in a Windows 11 Education version 23H2 with a processor Intel® Core^TM^ i7-3820 CPU @ 3.60GHz and 64 GB of RAM memory.

### 3.1. Datasets

For this study, we defined three datasets: the LEMON dataset, the AI-Mind dataset, and the test-retest dataset. These datasets include data from one public and one private source. Table 1 summarizes the number and types of EEG recordings in each dataset.

**Table 1.** Datasets used in the present study.

|  | N | Eyes closed | Eyes Open |
| --- | --- | --- | --- |
| LEMON dataset | 100 | 50 | 50 |
| AI-Mind dataset | 20 | 10 | 10 |
| Test-retest dataset | 20 | 10 pairs | 10 pairs |

#### 3.1.1. LEMON dataset

The “Leipzig Study for Mind-Body-Emotion Interactions” (LEMON)[16] is a publicly available dataset consisting of EEG recordings from 227 healthy participants comprising a young and an elderly group acquired cross-sectionally in Leipzig, Germany, between 2013 and 2015. For 216 of the participants, the creators recorded task-free (resting state) EEG using a BrainAmp MR plus amplifier in an electrically shielded and sound-attenuated EEG booth using a 62-channel (61 scalp electrodes plus 1 electrode recording the VEOG below the right eye) cap and active electrodes (actiCAP) (both Brain Products GmbH, Gilching, Germany) positioned according to the extended version or the international 10–20 system, (also known as 10-10 system), and referenced to FCz. The skin electrode impedance was kept below 5 KΩ. The EEGs were bandpass filtered online between 0.015 Hz and 1 kHz and digitized with a sampling rate of 2500 Hz. The EEG session included a total of 16 blocks, each 60 seconds long, 8 with eyes-closed (EC) and 8 with eyes-open (EO).

According to the database description, the raw EEG data was bandpass filtered between 1 and 45 Hz and downsampled to 250 Hz. Bad channels were rejected after visual inspection, looking for frequent jumps/shifts in voltage or poor signal quality. Data intervals containing extreme peak-to-peak deflections or large bursts of high frequency activity were identified by visual inspection and removed. Then, data was transferred to EEGLAB[17] (version 14.1.1b) for MATLAB. The dimensionality of the data was reduced using principal component analysis, by keeping the minimum number of components that explain 95% of the total data variance. Last, data was studied using the Infomax implementation (*runica*) of the independent component analysis[18] (ICA), and components reflecting eye movement, eye blinks, or heartbeat related artefacts were removed. Finally, data was segmented into 4-second epochs of clean data, plus 2 seconds of real data at each side, as padding.

For this study, we randomly selected 50 subjects resulting in 100 task-free EEG recordings (50 eyes-closed and 50 eyes-opened).

#### 3.1.2. AI-Mind dataset

The AI-Mind dataset [15] is a private dataset of 1022 participants within MCI continuum aged between 60 and 80 years, around 250 participants per each of the four involved countries (Spain, Italy, Norway, and Finland). For all participants, two 5-minutes eyes-closed EEG and two 5-minutes eyes-open EEG were recorded. EEGs were recorded using 126 electrodes prewired in an elastic cap (ANT neuro Waveguard™), in addition to two auxiliary electrodes placed below the left eye (electro-oculogram; EOG), and on the right clavicle bone (electrocardiogram; ECG). The electrodes were labelled according to the 10-5 system[19]. The impedances were kept as low as possible, preferably below 25 kΩ. The ground electrode was placed on the left mastoid, while the CPz electrode served as the online reference. The signals are amplified by an eego™ mylab EE-228 amplifier system and digitalized and stored using eego™ software, both provided by ANT neuro/eemagine Medical Imaging Solutions GmbH, Berlin, Germany. EEGs were recorded at a sampling rate of 2 kHz with an anti-aliasing filter with a high cut-off frequency of 520 Hz.

EEG recordings were manually preprocessed by four EEG experts from the Centre for Cognitive and Computational Neuroscience, including the first author (Federico Ramírez-Toraño). The expert team comprised two women and two men. Three experts hold PhDs in different disciplines (Biomedical Engineering, Physics, and Psychology), whereas the fourth expert is a PhD candidate in Biomedical Engineering. All experts have extensive experience in EEG preprocessing, as evidenced by their scientific contributions in the field.

The first step in the pipeline is a visual inspection for bad channels and artefacts. Spatially complex artefacts such as muscle activity might dominate the blind source separation, so segments containing these artefacts were excluded from this analysis. Then, data was studied using SOBI[20], and components associated to eye movements and heart beats were labelled and subtracted. After the component rejection, the data was again visually reviewed to confirm the artefacts present in the EEG recording. Finally, data was segmented into 4-second epochs of clean data, plus 2 seconds of real data at each side, as padding. The pipeline was coded in MATLAB, and it was based on Fieldtrip [14,21].

For this study, we randomly selected 20 subjects of the AI-Mind dataset. For each subject we selected one EEG recording, resulting in 20 task-free EEG recordings (10 eyes-closed and 10 eyes-opened).

#### 3.1.3. Test-retest dataset

To perform consistency analysis, we created another dataset from a subset of the AI-Mind dataset. Specifically, from the AI-Mind dataset, another 20 subjects were randomly selected. Each subject has 4 EEG recordings (two eyes-closed resting and two eyes-opened) obtained during the same session in an interleaved fashion (eyes-open, eyes-closed, eyes-open, eyes-closed).

### 3.2. Performance measurements

#### 3.2.1. Metadata

For each EEG recording and each of the conceptual modules defined in section 2, we stored the time used to perform the task and the peak memory consumed during the task. For sEEGnal we used the Python modules *time* and *tracemalloc*, respectively. For the EEG experts, we used the MATLAB functions *tic-toc* and *memory*, respectively.

In addition, we stored the number of bad channels, the total duration of artefacts, and ICs rejected.

#### 3.2.2. Power spectrum

For each channel, the power spectrum was calculated by means of Welch’s method using a Hann window of 4 seconds (this is, after removing the padding) and 0.25 Hz of spectral resolution. This method estimated the periodogram between 2 and 45 Hz for each epoch and then averaged across epochs. The relative power spectrum of each channel was obtained by normalizing the power spectrum by the total broadband power, resulting in a value ranging from 0 to 1. As a result, a 2D matrix (*number of channels × frequency steps*) was obtained for each recording and pre-processing approach.

#### 3.2.3. Functional connectivity

Functional connectivity was estimated using the phase locking value (PLV)[22,23] for each classical frequency band (delta, theta, alpha, low beta, high beta, gamma)[24]. This measure is based on the hypothesis of *phase synchronization* for connected brain activity, i.e. that the instantaneous phase of two connected signals should evolve together, and, therefore, the difference between them should be bounded. According to how “locked” this difference is through the EEG epoch, the PLV ranges from 0 to 1, where the lower bound indicates no connectivity and the upper full connectivity.

For each epoch, the signal was filtered in one of the classical bands, the padding was removed, and the PLV was estimated for each pair of channels. Finally, the PLV for the pair of channels was estimated as the average across epochs. As result, a 3D matrix (*number of channels × number of channels × frequency band*) was obtained for each recording and pre-processing approach.

### 3.3. Statistical analysis

The analysis performed in this study intend to evaluate two main aspects of the proposed solution: the performance of sEEGnal pipeline compared to a human-driven pipeline, and the consistency of the result between two recordings within the same session. A global overview of the statistical analysis is presented in Figure 4.

**Figure 4.**
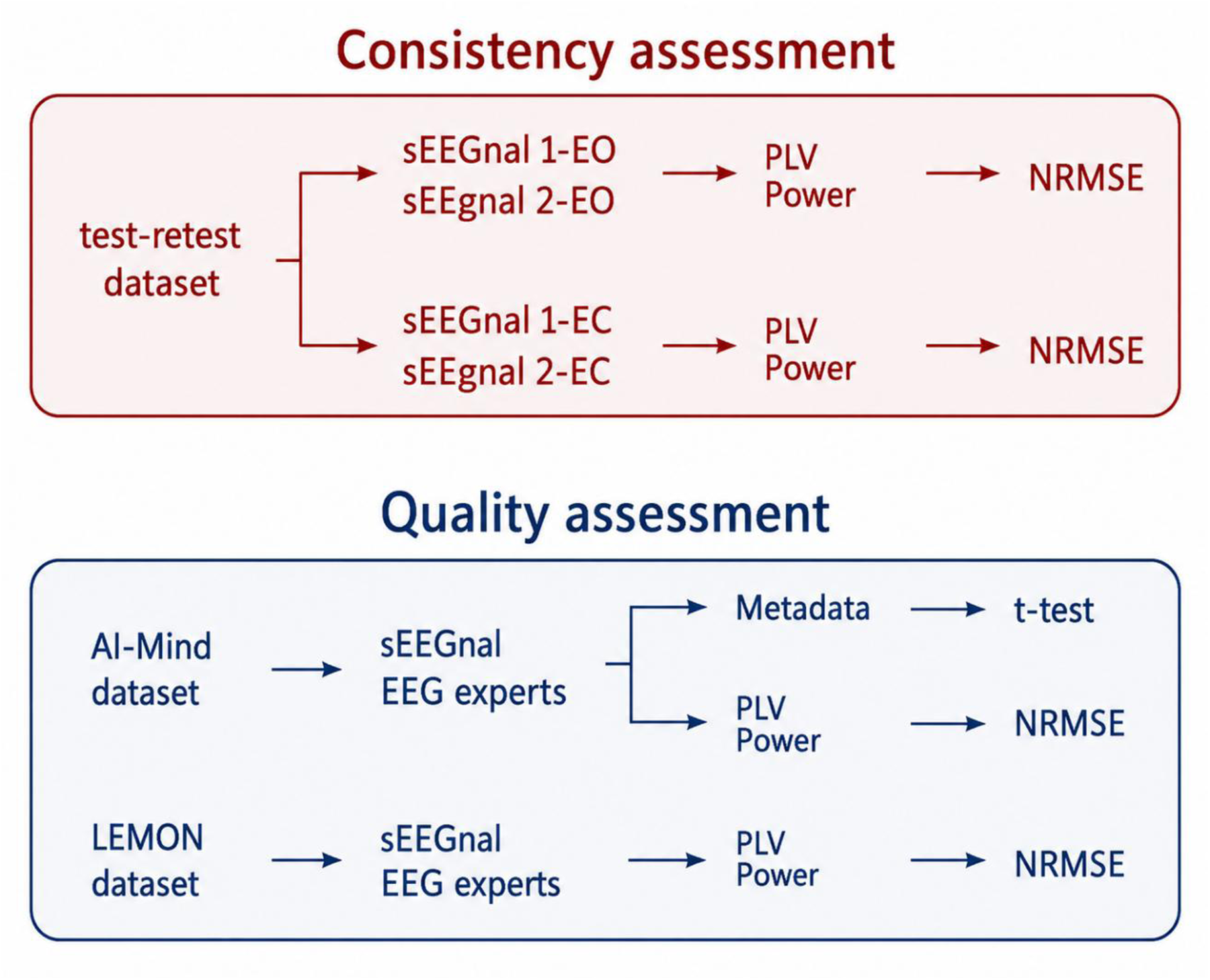
Global map of the analysis performed in this study. Abbreviations: PLV, phase locking value; NRMSE, normalised root mean squared error.

We compared the metadata produced by sEEGnal and by human experts in the AI-Mind dataset to evaluate whether the preprocessing decisions made by sEEGnal were comparable to expert-driven decisions. First, we assessed differences in processing time, memory consumption, number of bad channels detected, total duration of detected artefacts (pooled across all artefact categories), and number of rejected ICs between sEEGnal and the EEG experts using paired t-tests.

Focusing on bad channels, we evaluated their spatial distribution across sensors and performed Bland–Altman and Dice overlap analyses to assess agreement between sEEGnal and EEG experts.

Focusing on artefacts, we analysed the distribution of artefact categories (EOG, muscle, jump, and other) identified by sEEGnal and EEG experts, and we performed Bland–Altman and temporal Dice overlap analyses. For the temporal Dice analysis, artefact annotations were converted into binary temporal masks using a temporal resolution of 0.1 seconds.

Dice overlap was then estimated as 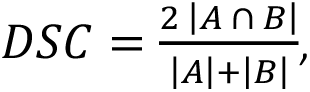 where (A) and (B) corresponded to the temporal masks generated by two preprocessors. Cases where both preprocessors did not identify any artefact of a given category (Dice = 1 by definition) were excluded from the analysis to avoid artificially inflating agreement estimates.

Focusing on rejected ICs, we compared the number of ICs rejected by sEEGnal and EEG experts and performed Bland–Altman analyses to assess agreement.

We assessed the quality of sEEGnal through the output measurements (power spectra and functional connectivity) using the normalized root mean squared error (NRMSE). NRMSE offers a dimensionless estimation of similarity between two measures:

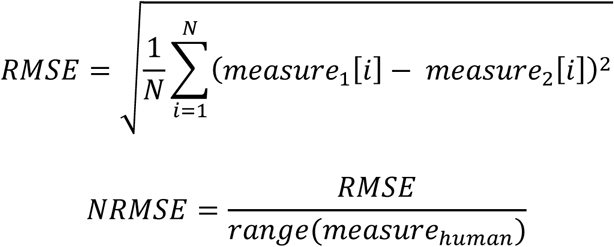

An NRMSE close to 0 indicates a high degree of similarity between the signals, while NRMSE close to 1 suggest differences comparable to the total magnitude of the signal.

To assess the consistency of sEEGnal, we estimated the NRMSE values for same-session EEG recordings from the Test-retest dataset. Specifically, for each subject and session, we compared the eyes-closed recordings and the eyes-opened task-free recordings separately.

To assess the performance of sEEGnal, we estimated the NRMSE between EEG recording from the AI-Mind dataset and LEMON dataset pre-processed either with sEEGnal or by EEG experts. Finally, to gain a comprehensive insight and detect any possible spatial or frequency bias, the NRMSE values were plotted for each sensor and frequency band.

For example, to compare the power spectrum after pre-processing by sEEGnal and an EEG expert, first, we loaded the power spectrum obtained from all the EEG recordings with the two pre-processing approaches. Second, we selected, from each recording, the desired frequency band and channel. Third, we estimated the NRMSE between the values for the specific frequency band and sensor pre-processed by sEEGnal and the EEG expert. Finally, to obtain a visual insight of the spatial “quality map” the NRMSE values were averaged across recordings (Figure 5). For the sake of clarity, we present a pseudocode outlining the method in the Supplementary Material.

**Figure 5.**
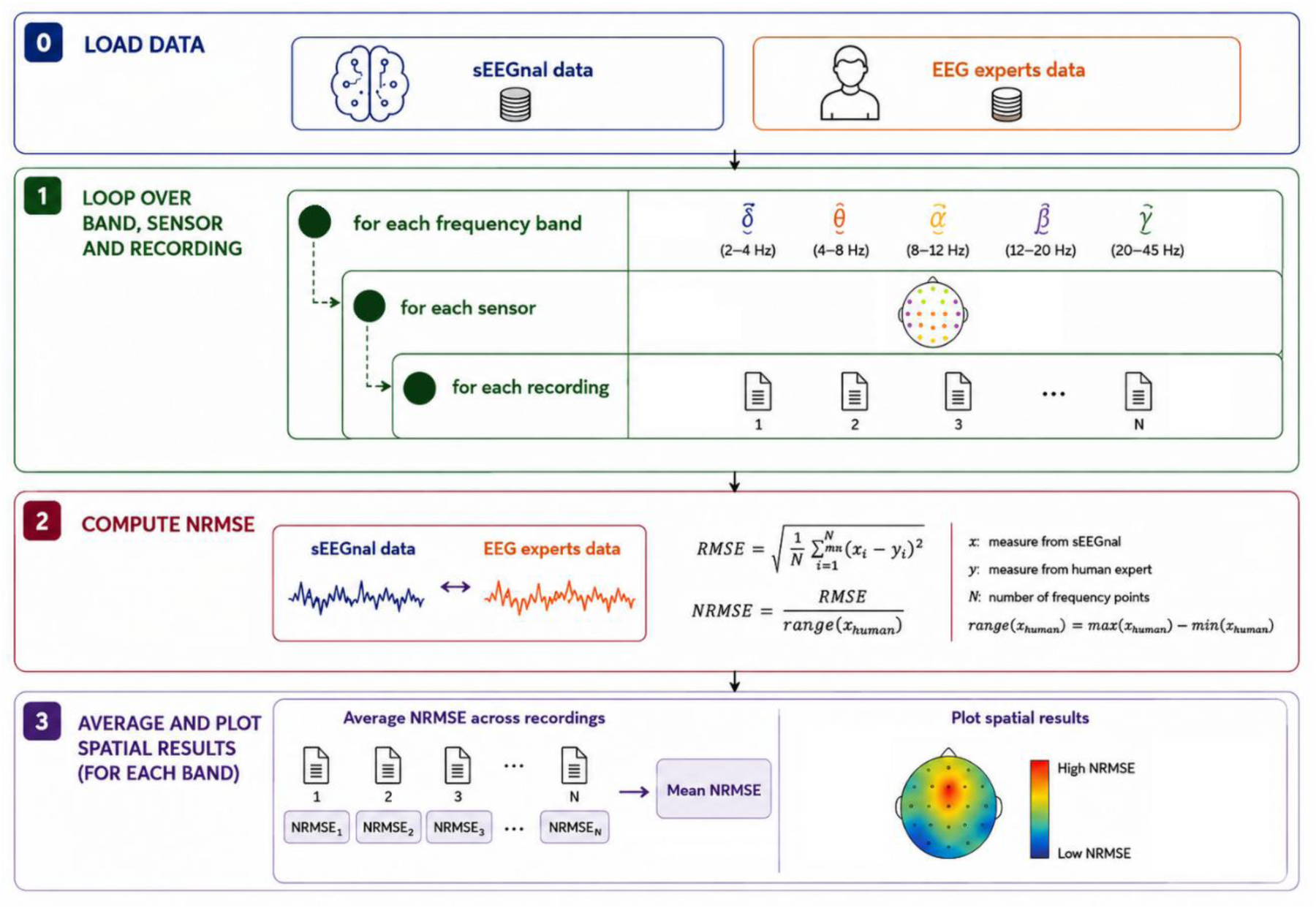
Pipeline to assess sEEGnal performance using the power spectrum. For the sake of clarity, this example is simplified by considering only two recordings, the frontal area, and one metric. The power spectra of frontal channels are estimated using the data obtained either by sEEGnal o f the human EEG experts. The power spectra of each channel are compared using correlation and NRMSE, and the average is plot in head space for spatial visualization.

## 4. Results

### 4.1. Metadata

#### 4.1.1. AI-Mind dataset

Table 2 presents the metadata results. Regarding time spent by each module, we observed similar average time in each conceptual module but greater standard deviation in the EEG experts. We observed less memory consumed by the proposed pipeline sEEGnal.

**Table 2.** Metadata measures regarding performance for EEG experts and for sEEGnal. Time is measured in seconds, Memory in Megabytes. The results are presented as mean (standard deviation). Due to a human mistake, the metadata regarding EEG expert memory in artefact detection is lost.

| Measure | Module | sEEGnal | EEG experts | p-value | Cohen's d |
| --- | --- | --- | --- | --- | --- |
| Time | Bad channel detection | 324.40<br>(85.45) | 372.55<br>(561.65) | 0.710 | 0.173 |
|  | Artefact detection | 373.52<br>(91.30) | 425.70<br>(235.40) | 0.318 | 0.470 |
| Memory | Bad channel detection | 2473.82<br>(620.89) | 3134.19<br>(259.83) | < 0.001 | 1.911 |
|  | Artefact detection | 2399.50<br>(602.38) | - | - | - |
| Number of bad channels |  | 6.05<br>(3.41) | 6.21<br>(3.59) | 0.863 | 0.080 |
| Artefacts in seconds |  | 16.71<br>(17.61) | 20.24<br>(19.48) | 0.172 | -0.650 |
| Number of ICs rejected |  | 2.10<br>(1.12) | 1.82<br>(0.70) | 0.239 | -0.557 |

Regarding the number of bad channels, artefacts, and components rejected, we observed a similar performance with both pipelines.

The spatial distribution of the bad channels is presented in Figure 6. For clarity purposes, the channel distribution was divided into groups according to the percentage of times the bad channel was marked as bad (from marked as bad in 45% of the recordings to 0 times marked as bad). First, we observed that the maximum proportion of recordings in which a channel was marked as bad reached 45% for sEEGnal, whereas it was 16% for human experts. The channels most frequently identified as bad by sEEGnal were predominantly located in frontal and temporal regions, a spatial pattern that was also observed in expert annotations.

**Figure 6.**
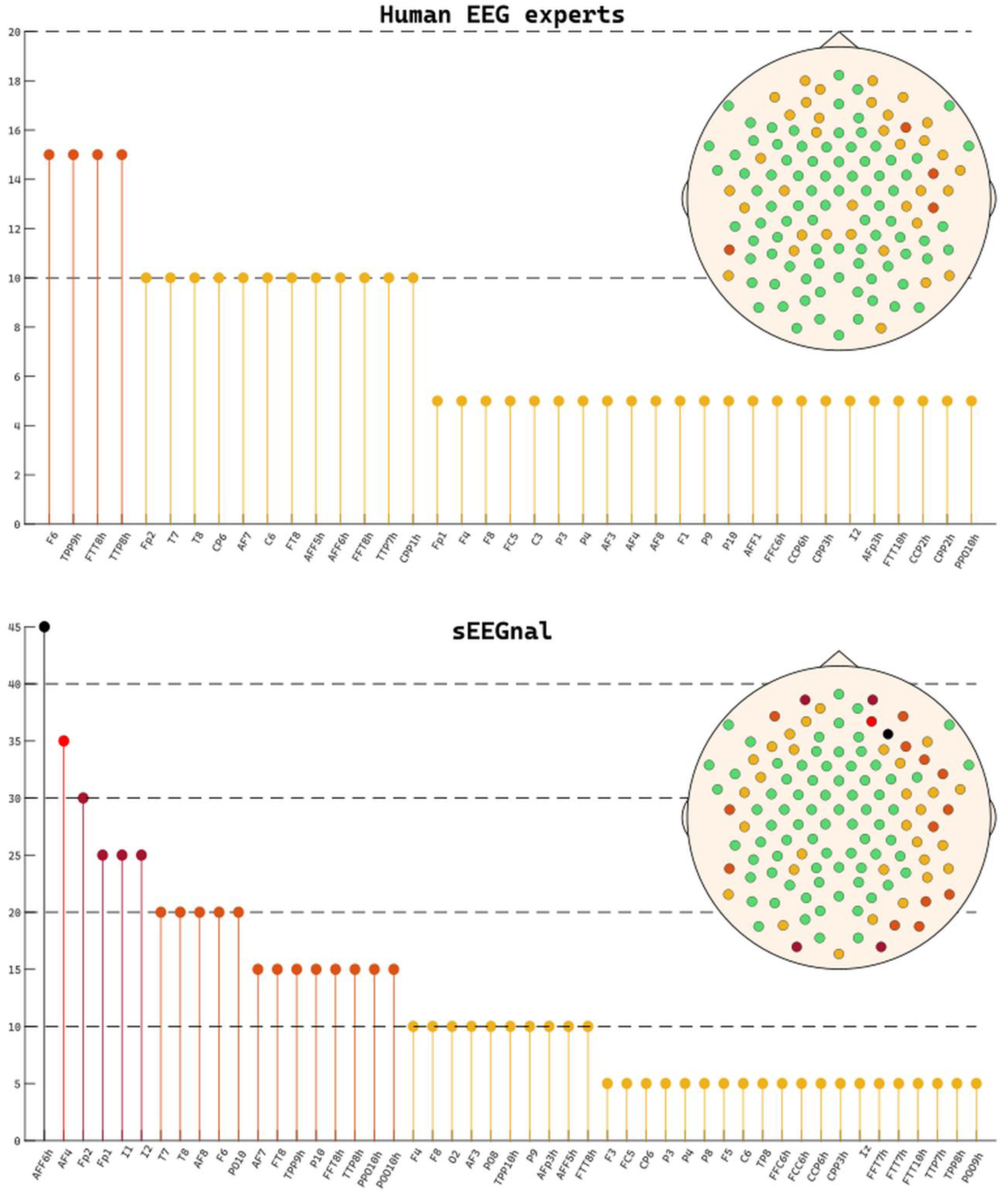
Spatial distribution of bad channels. On the top panel the information regarding the EEG experts. On the bottom panel the information regarding sEEGnal. The colormap divides the bad channels in groups according to their incidence in the EEG recordings pre-processed. For clarity purposes, the channel distribution was divided into groups according to the percentage of times the bad channel was marked as bad (from marked as bad in 45% of the recordings to 0 times marked as bad)

The Bland–Altman analysis revealed minimal systematic bias (bias = −0.16 channels) and moderate variability across recordings (Limts of Agreement (LoA): [−8.31, 7.99]).

Importantly, this variability was comparable to the inter-expert variability observed between human EEG experts (Figure 7).

**Figure 7.**
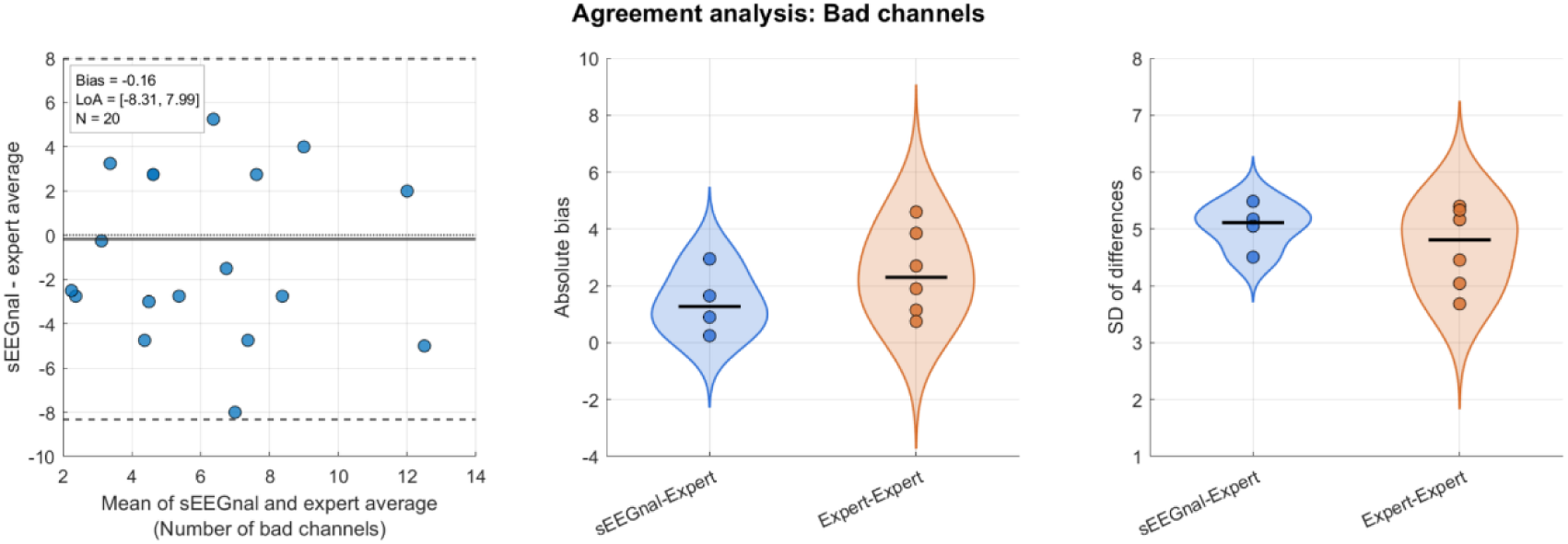
Agreement analysis for bad channel detection. Left: Bland –Altman plot comparing the number of bad channels identified by sEEGnal and the average expert annotation across recordings. The solid horizontal line indicates the mean bias, and dashed lines indicate the limits of agreement (LoA). Middle: Distribution of the absolute bias for sEEGnal–expert and expert–expert comparisons. Right: Distribution of the standard deviation of differences for sEEGnal –expert and expert–expert comparisons.

The Dice overlap analysis showed that the spatial agreement in bad channel identification between sEEGnal and EEG experts was comparable to the agreement observed between pairs of human experts (Figure 8).

**Figure 8.**
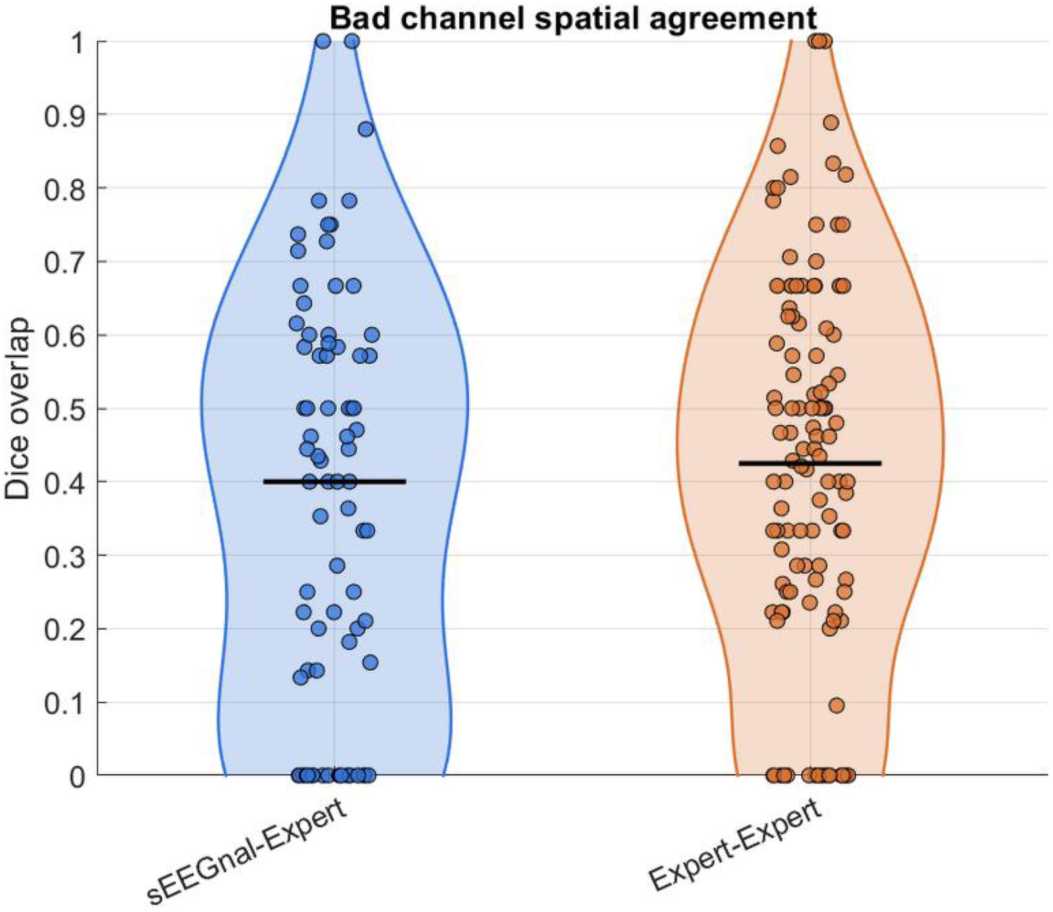
Spatial agreement analysis for bad channel detection. Violin plots represent the distribution of Dice overlap values for sEEGnal –expert and expert–expert comparisons across recordings. Each point corresponds to a single comparison, and horizontal black l ines indicate the median Dice overlap.

The types of ICs and the types of artefacts are presented in Figure 9. Regarding the ICs, 1.59 % of the components were labelled for rejection by EEG experts, while 7.14 % were labelled for rejection by sEEGnal. Regarding the types of artefacts, EEG experts classified 32.82 % of the artefacts as eye-related, 21.02 % as muscle-related, 39.85 % as electronic jumps, and 6.32 % as “other”. For sEEGnal, 44.01 % of the artefacts were eye-related, 36.94 % muscle-related, 17.29 % electronic jumps, and 1.76 % “other”. A chi-square test of homogeneity revealed significant differences in the distribution of artefact types between sEEGnal and EEG experts (χ² (3) = 124.57, p < 0.001).

**Figure 9.**
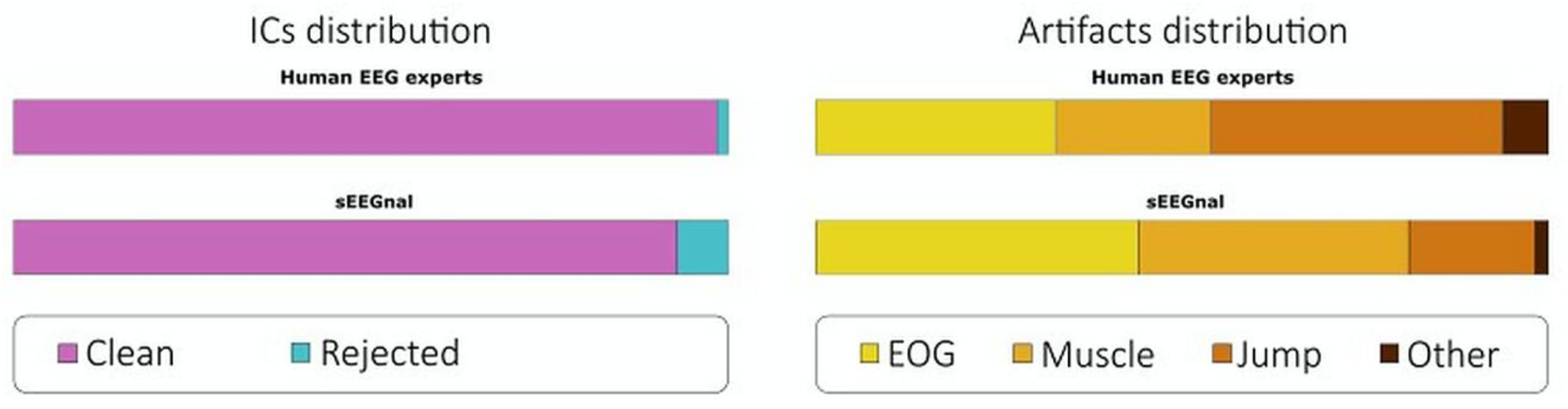
Details of the ICs and artefacts present in the recordings. On the left, the distribution of ICs according to their type. On the right, the types of artefacts. Abbreviations: EOG, electrooculogram.

For the number of artefacts, Bland–Altman analysis revealed a moderate positive bias (bias = 6.06 artefacts) and relatively broad limits of agreement (LoA = [−31.42, 43.55]). Importantly, the agreement comparison analyses showed that the values observed between sEEGnal and human experts was comparable to the values observed between pairs of human experts (Figure 10).

**Figure 10.**
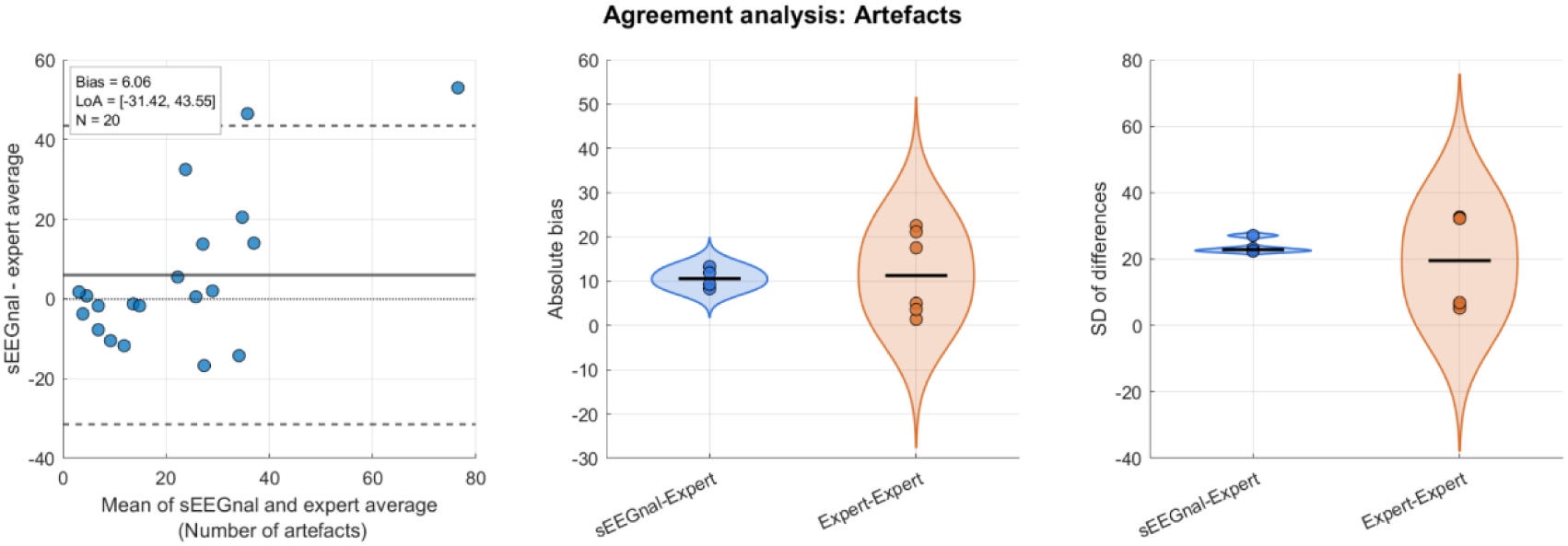
Agreement analysis for artefacts detection. Left: Bland –Altman plot comparing the number of artefacts identified by sEEGnal and the average expert annotation across recordings. The solid horizontal line indicates the mean bias, and dashed lines indicate the limits of agreement (LoA). Middle: Distribution of the absolute bias for sEEGnal–expert and expert–expert comparisons. Right: Distribution of the standard deviation of differences for sEEGnal –expert and expert–expert comparisons.

The temporal Dice overlap analysis revealed differences across artefact categories. EOG and “other” artefacts showed overall low temporal Dice overlap values for both sEEGnal–expert and expert–expert comparisons. Jump artefacts showed moderate temporal overlap, with slightly higher Dice values for expert–expert comparisons. Muscle artefacts showed the highest temporal Dice overlap values overall, particularly for expert–expert comparisons (Figure 11).

**Figure 11.**
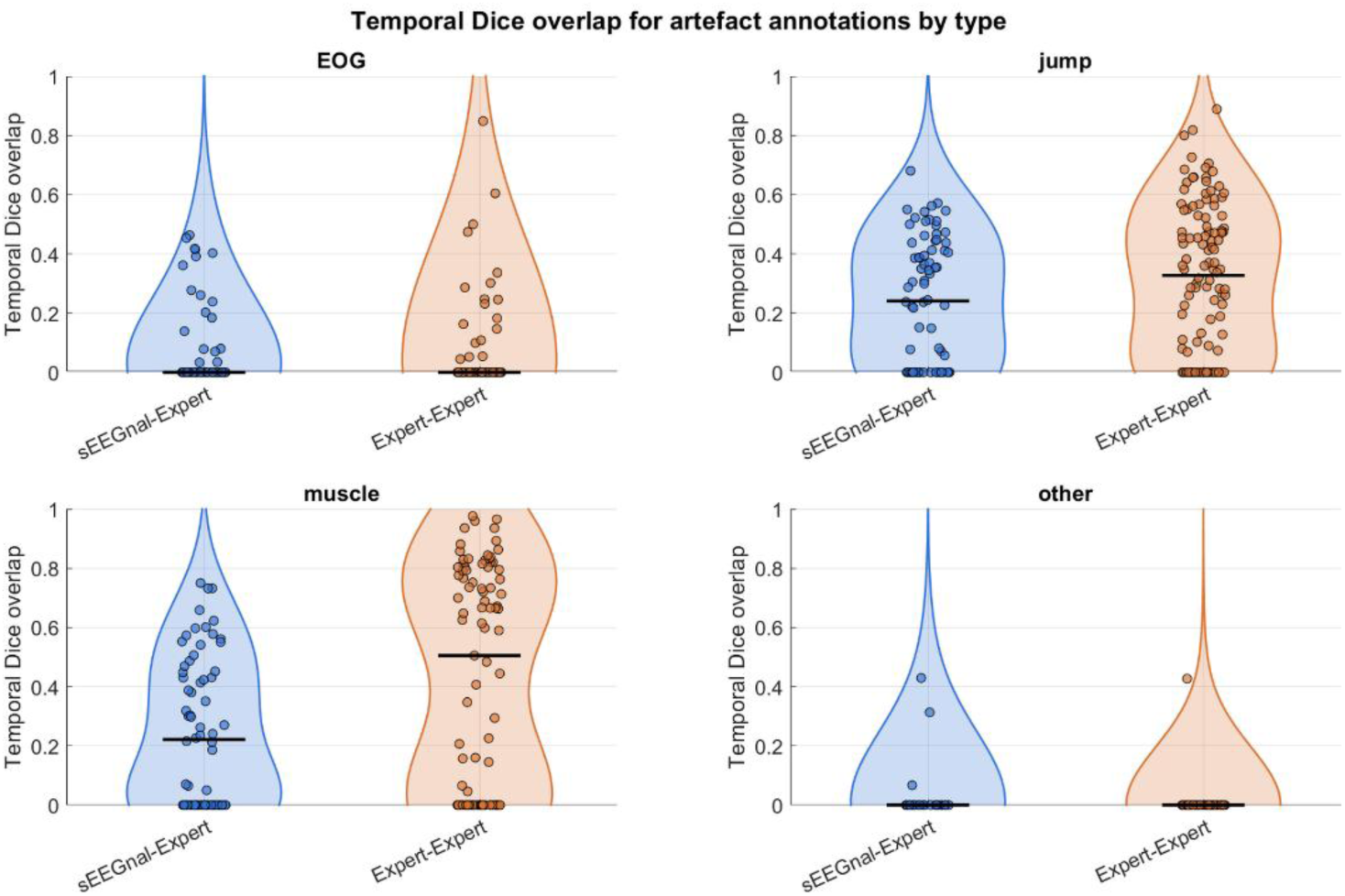
Temporal agreement analysis for artefacts detection. Violin plots represent the distribution of Dice overlap values for sEEGnal –expert and expert–expert comparisons across recordings. Each point corresponds to a single comparison, and horizontal black lines indicate the median Dice overlap.

For the number of ICs rejected, the Bland–Altman analysis revealed minimal systematic bias between sEEGnal and the average expert annotation (bias = 0.28 ICs; LoA = [−1.71, 2.26]). The agreement comparison analyses showed similar distributions of absolute bias and standard deviation of differences for sEEGnal–expert and expert–expert comparisons (Figure 12).

**Figure 12.**
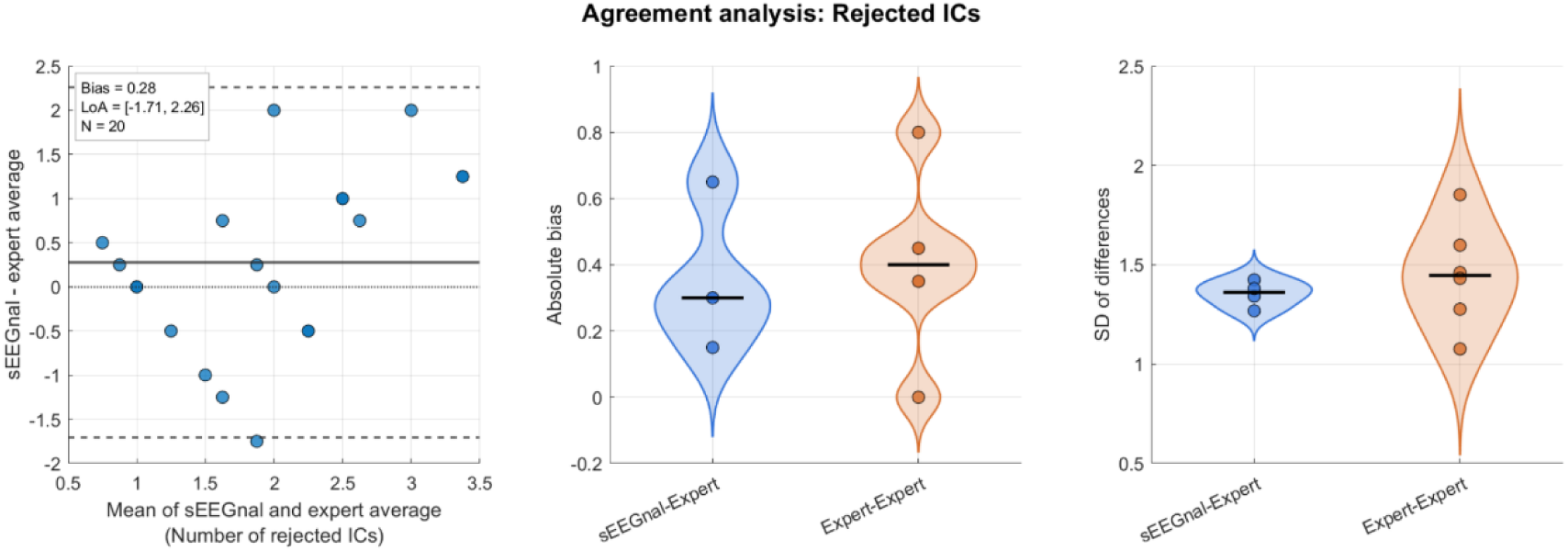
Agreement analysis for ICs rejected. Left: Bland –Altman plot comparing the number of artefacts identified by sEEGnal and the average expert annotation across recordings. The solid horizontal line indicates the mean bias, and dashed lines indicate the limits of agreement (LoA). Middle: Distribution of the absolute bias for sEEGnal–expert and expert–expert comparisons. Right: Distribution of the standard deviation of differences for sEEGnal –expert and expert–expert comparisons.

The complete Bland-Altman analysis, i.e. one to one comparison among sEEGnal and all human experts and among human experts, can be found in Supplementary Material.

### 4.2. Power spectra

#### 4.2.1. LEMON dataset

The power spectra obtained by sEEGnal showed a great resemblance to the ones averaged from the EEG experts (as an example, see Figure 13). For purposes of visualization, the power spectra obtained by both sEEGnal and the EEG experts are plotted in Figure 13 (top panel). The global resemblance is presented in Figure 13 (bottom panel), where the power of each frequency band obtained by sEEGnal for all subjects and channels are plotted against the ones obtained by the EEG experts. The correlation between sEEGnal and human experts was significant for all bands (p < 0.0001) with the lowest correlation value observed in delta band (rho = 0.8624).

**Figure 13.**
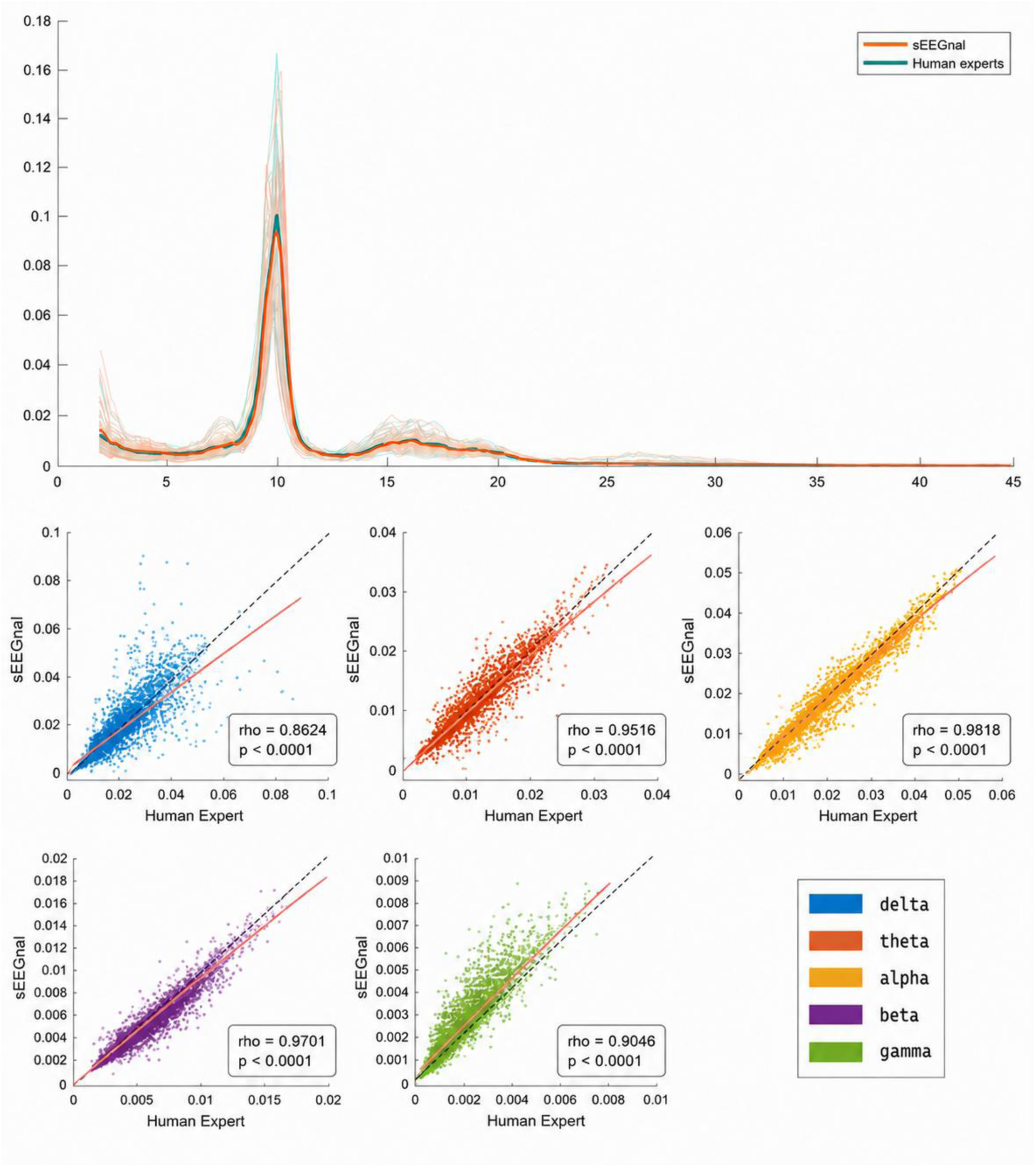
Comparison of power spectra obtained by the EEG experts and by sEEGnal in the LEMON dataset. On the top panel, the power spectra of a subject processed by sEEGnal and by EEG experts. The pale lines represent individual channels, and the solid lines represent the average power spectrum across channels for each preprocessing approach. On the bottom panel, an X-Y plot of the value obtained by sEEGnal and EEG expert dataset for the power in each band. The dash line represents a perfect correspondence (1:1), a nd the pink line represents the least-squares line.

Regarding the frequency bands, the NRMSE values for all channels in all recordings analysed are below 0.08, meaning the maximum difference across all the data analysed is below 8 % of the metric range, with the delta band being the one with larger differences.

(Figure 14, top). Regarding the spatial distribution of the differences, the frontal channels in the delta band presented the highest differences, followed by the alpha band widespread across the head (Figure 14, bottom panel).

**Figure 14.**
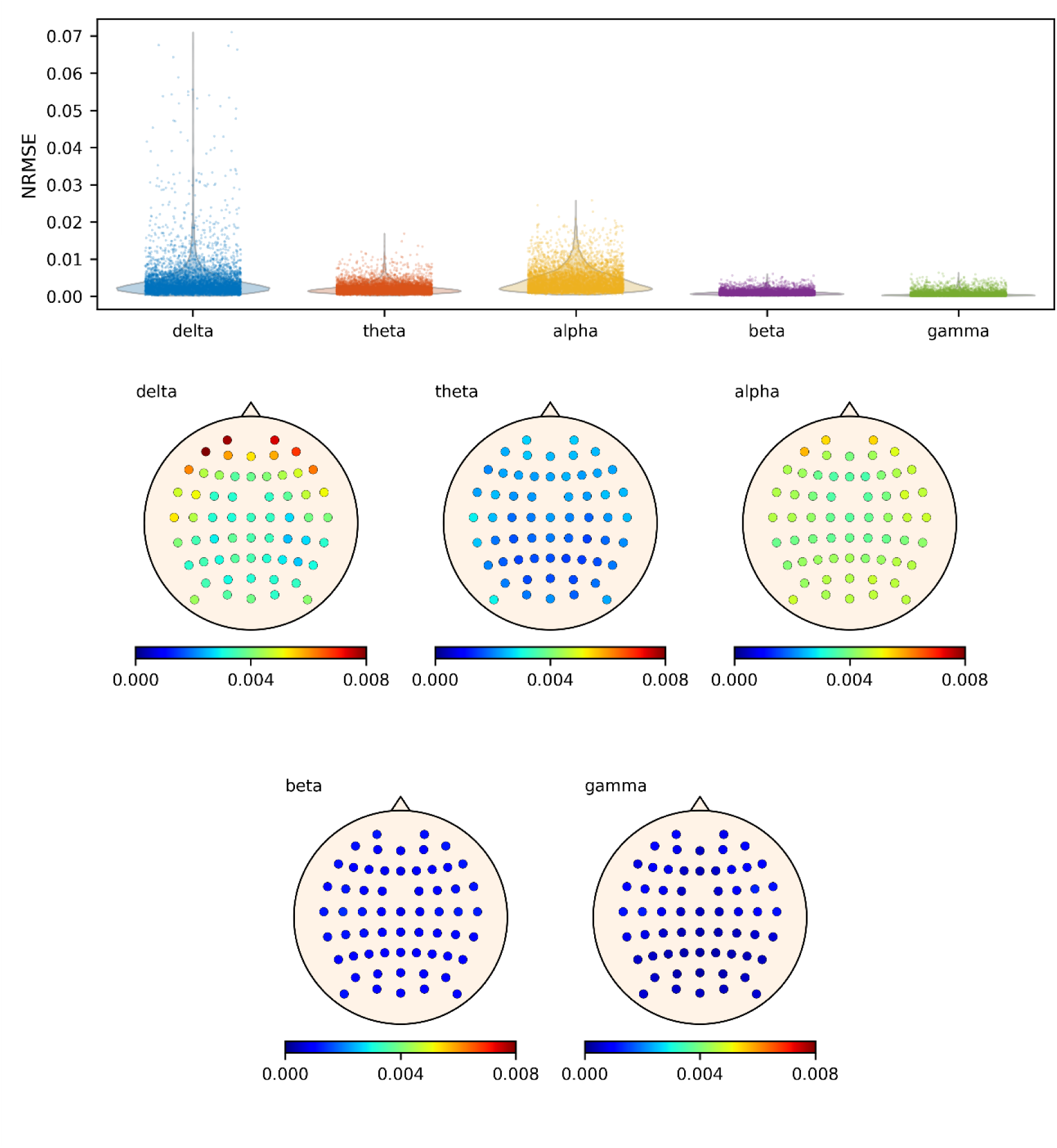
NRMSE values of the power spectra in the LEMON dataset. On the top panel, violin plots representing of the NRMSE distributions for all channels for each frequency band. On the bottom panels, spatial distribution of NRMSE values across the head for each frequency band. Abbreviations: NRMSE, normalised root mean square error.

#### 4.2.2. AI-Mind dataset

The power spectra obtained by sEEGnal showed high resemblance to the ones averaged from the EEG experts (Figure 15). For purposes of visualization, the power spectra obtained by sEEGnal and the EEG experts are depicted in Figure 15 (top panel). The global resemblance is presented in Figure 15 (bottom panel), where the average power of each frequency band obtained by sEEGnal for all subjects and channels are plotted against the ones obtained by the EEG experts in an X-Y plot. The correlation between sEEGnal and human experts was significant for all bands (p < 0.0001) with the lowest correlation value observed in delta band (rho = 0.8034).

**Figure 15.**
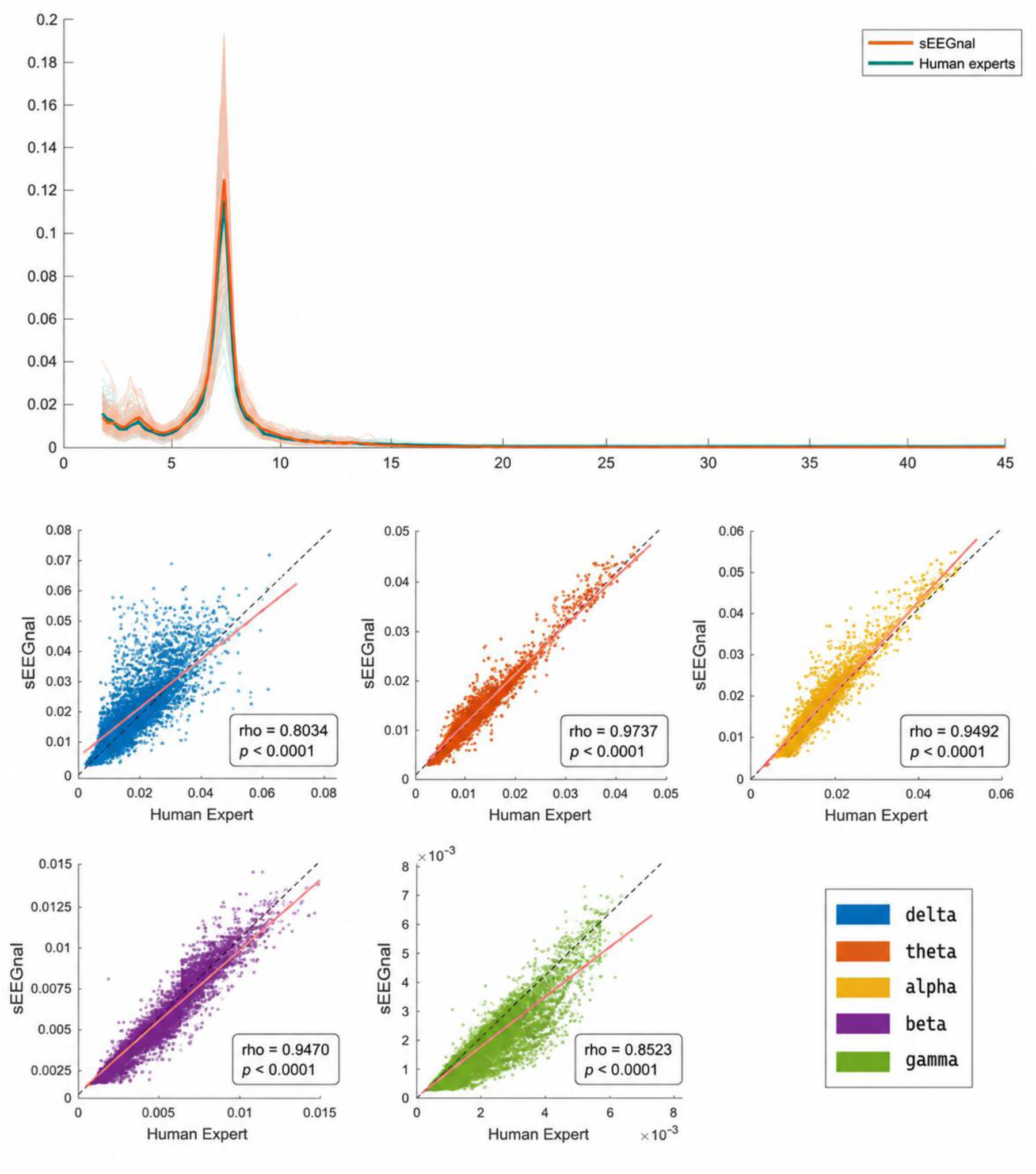
Comparison of power spectra obtained by the EEG experts and by sEEGnal in the AI-Mind dataset. On the top panel, the power spectra of a subject processed by sEEGnal and by EEG experts. The pale lines represent individual channels, and the solid lines represent the average power spectrum across channels for each preprocessing approach. On the bottom panel, an X-Y plot of the value obtained by sEEGnal and EEG expert dataset for the power in each band. The dash line represents a perfect correspondence (1:1), and the pink line represents the least-squares line.

Regarding the power per frequency band, the NRMSE values for all channels in all recordings analysed are below 0.08, meaning the maximum difference across all the data analysed is below 8 % of the metric range, with delta band being the one with more differences. (Figure 16, top). Regarding the spatial distribution of the differences, all channels present NRMSE values below 1.6 % of the range of the metric (Figure 16, bottom panel), with the frontal channels in the delta band presenting the largest differences, followed, as in the case of the LEMON dataset, for the alpha band.

**Figure 16.**
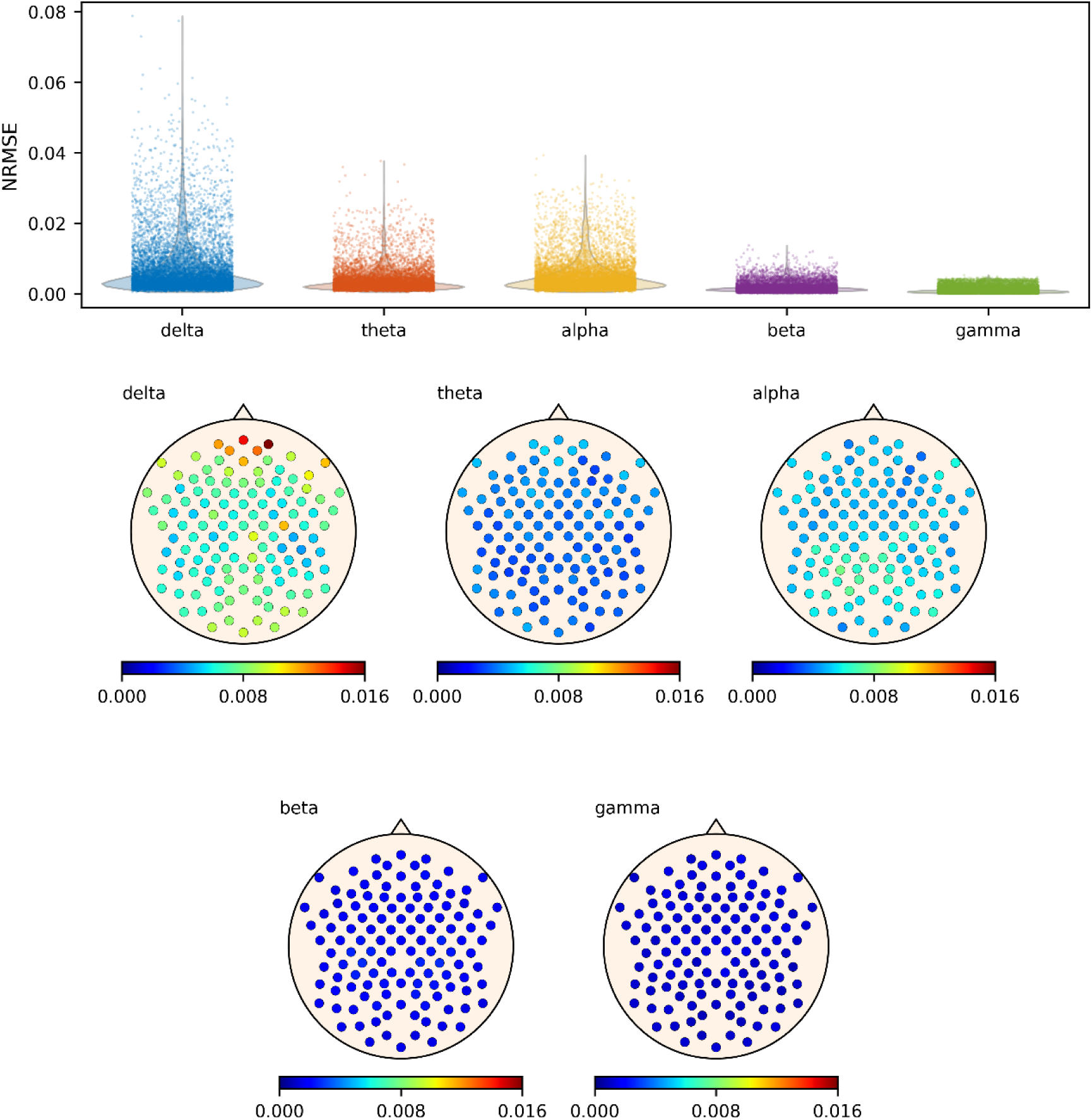
NRMSE values of the power spectra in the AI-Mind dataset. On the top panel, violin plots representing of the NRMSE distributions for all channels for each frequency band. On the bottom panels, spatial distribution of NRMSE values across the head for each frequency band. Abbreviations: NRMSE, normalised root mean square error.

#### 4.2.3. Test-retest dataset

For the sake of clarity, only the results for eyes-closed task-free data are presented in this section, with those for eyes-open presented in the Supplementary Material.

The power spectra obtained from the first and second recording of the same session are presented in Figure 17 (top panel). The global resemblance is presented in Figure 17 (bottom panel), where the average power of each frequency band obtained by sEEGnal for all first recordings are plotted against the ones obtained for all second recordings in an X-Y plot. The correlation between the first and the second recording was significant for all bands (p < 0.0001) with the lowest correlation value observed in gamma band (rho = 0.8668).

**Figure 17.**
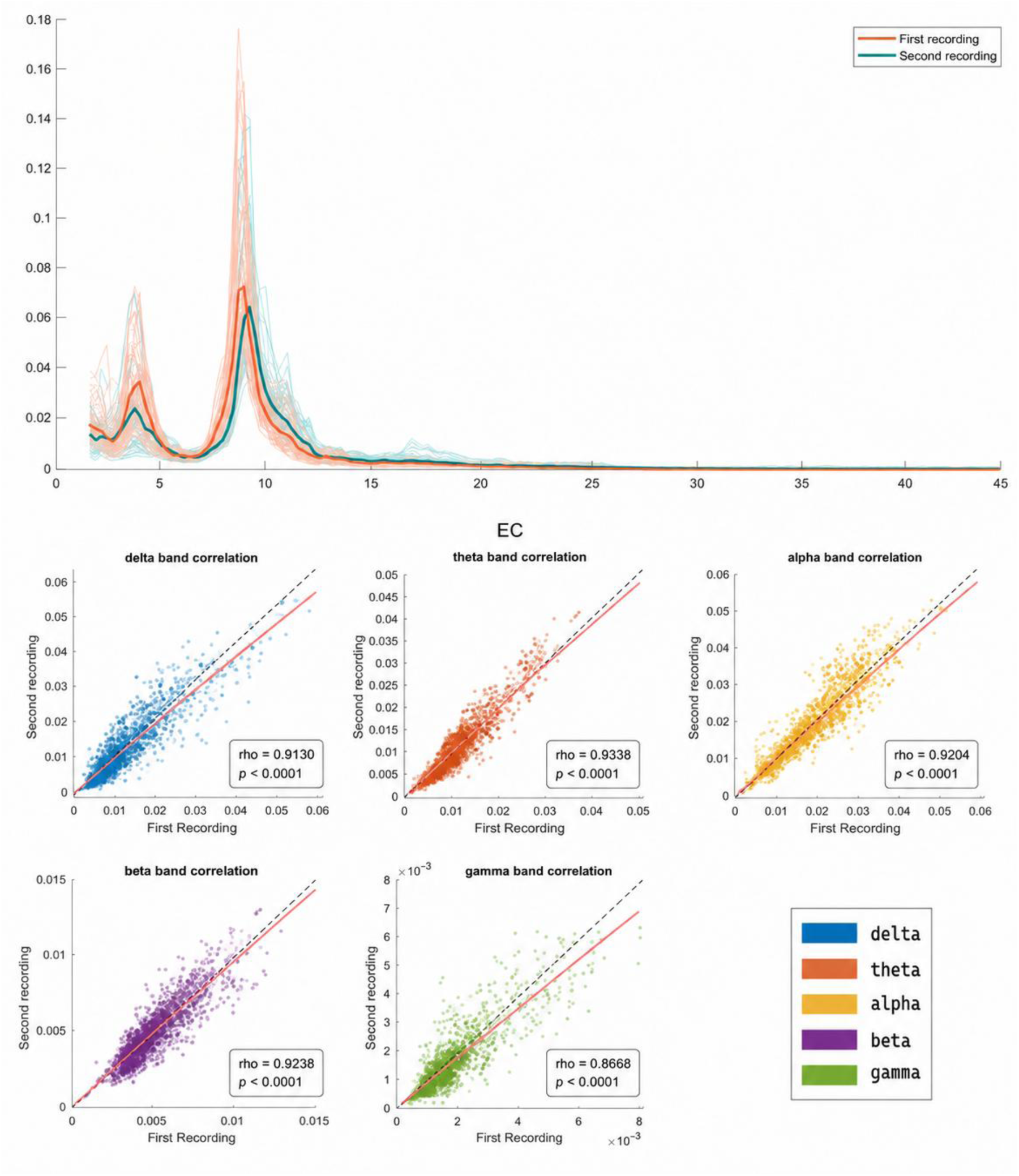
Comparison of power spectra obtained by sEEGnal on the test-retest dataset. On the top panel, the power spectra of both recordings. The pale lines represent individual channels, and the solid lines represent the average power spectrum across channels. On the bottom panel, an X-Y plot of the value obtained by sEEGnal on both recordings for the power in each band. The dash line represents a perfect correspondence (1:1), and the pink line represents the least-squares line.

Regarding the results per frequency band, the NRMSE values for all channels in all recordings analysed are below 0.07, meaning the maximum difference across all the data analysed is below 7 % of the metric range, with alpha band being the one with largest differences. (Figure 18, top panel). Regarding the spatial distribution of the differences, all channels present NRMSE values below 0.011 (1.1 % of the range of the metric) (Figure 18, bottom panel), with the occipital channels in alpha band presenting the largest differences.

**Figure 18.**
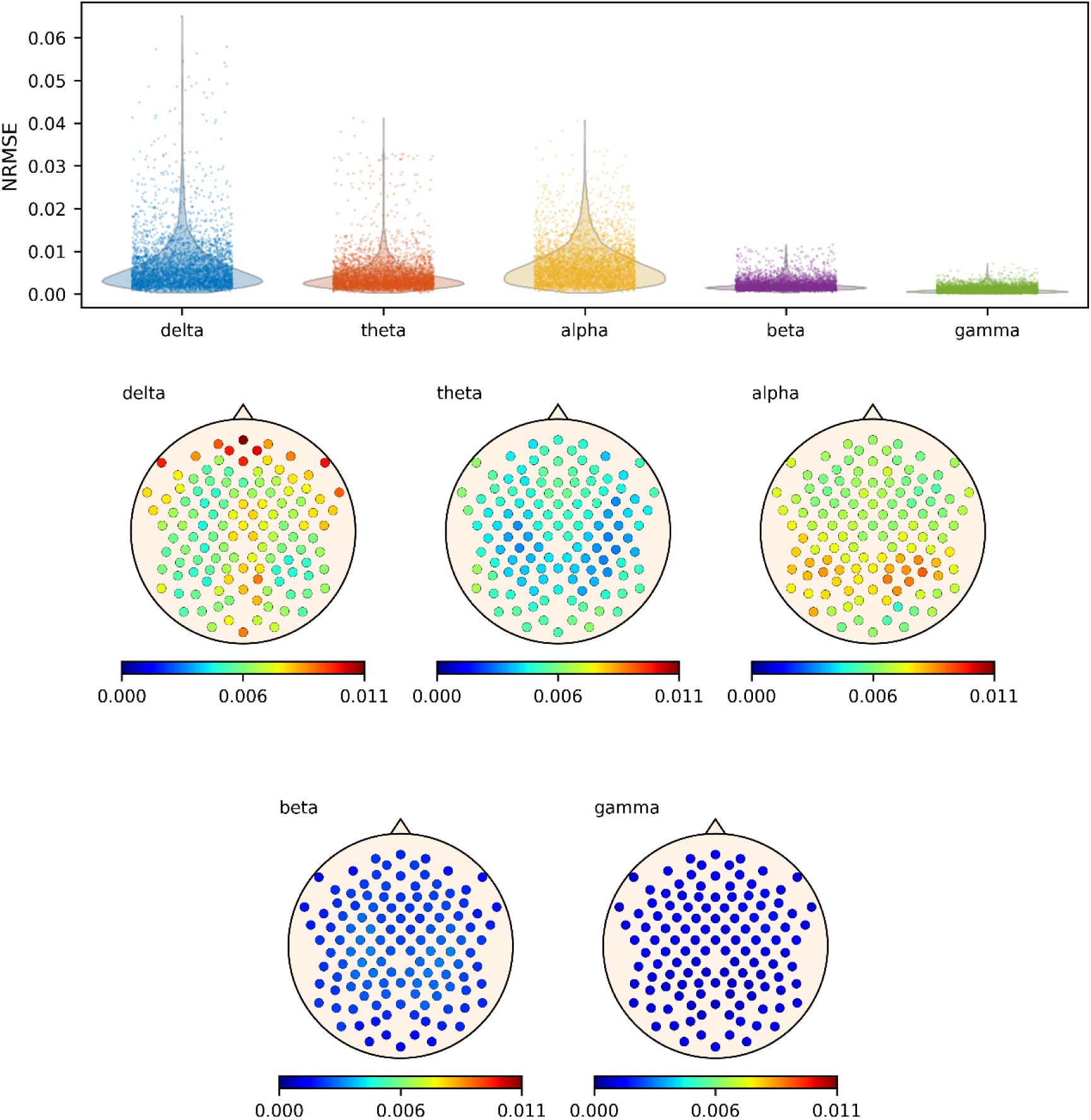
NRMSE values of the power spectra in the test-retest dataset. On the top panel, violin plots representing of the NRMSE distributions for all channels for each frequency band. On the bottom panels, spatial distribution of NRMSE values across the head for each frequency band. Abbreviations: NRMSE, normalised root mean square error.

### 4.3. Functional Connectivity

#### 4.3.1. LEMON dataset

The PLV obtained by sEEGnal showed a great resemblance to the ones averaged from the EEG experts (Figure 19). The global resemblance is presented as the PLV of each frequency band obtained by sEEGnal for all subjects and channels against the ones obtained by the EEG experts. The correlation between sEEGnal and human experts was significant for all bands (p < 0.0001) with the lowest correlation value observed in gamma band (rho = 0.9238).

**Figure 19.**
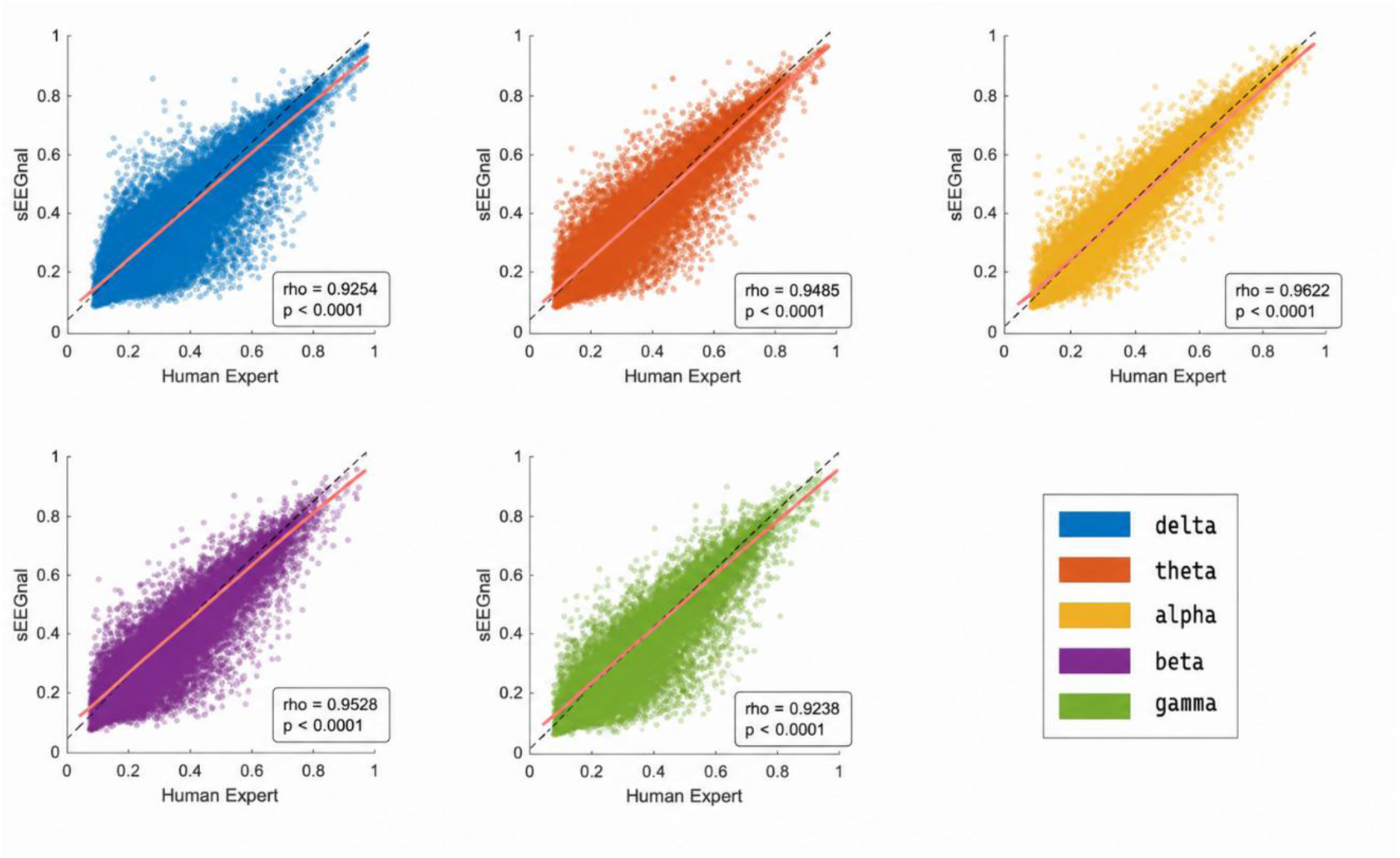
Comparison of PLV values obtained by the EEG experts and by sEEGnal in the LEMON dataset. An X-Y plot of the PLV values obtained in each band. The dash line represents a perfect correspondence (1:1), and the pink line represents the least-squares line.

Regarding the results per frequency band, the NRMSE values for all channels in all recordings analysed are below 0.17, meaning the maximum difference across all the data analysed is below 17 % of the metric range, with gamma band being the one with more differences. (Figure 20, top). Regarding the spatial distribution of the differences, all channels present NRMSE values below 5 % of the range of the metric (Figure 20, bottom panel), with the frontal channels presenting the highest differences, especially in the gamma band.

**Figure 20.**
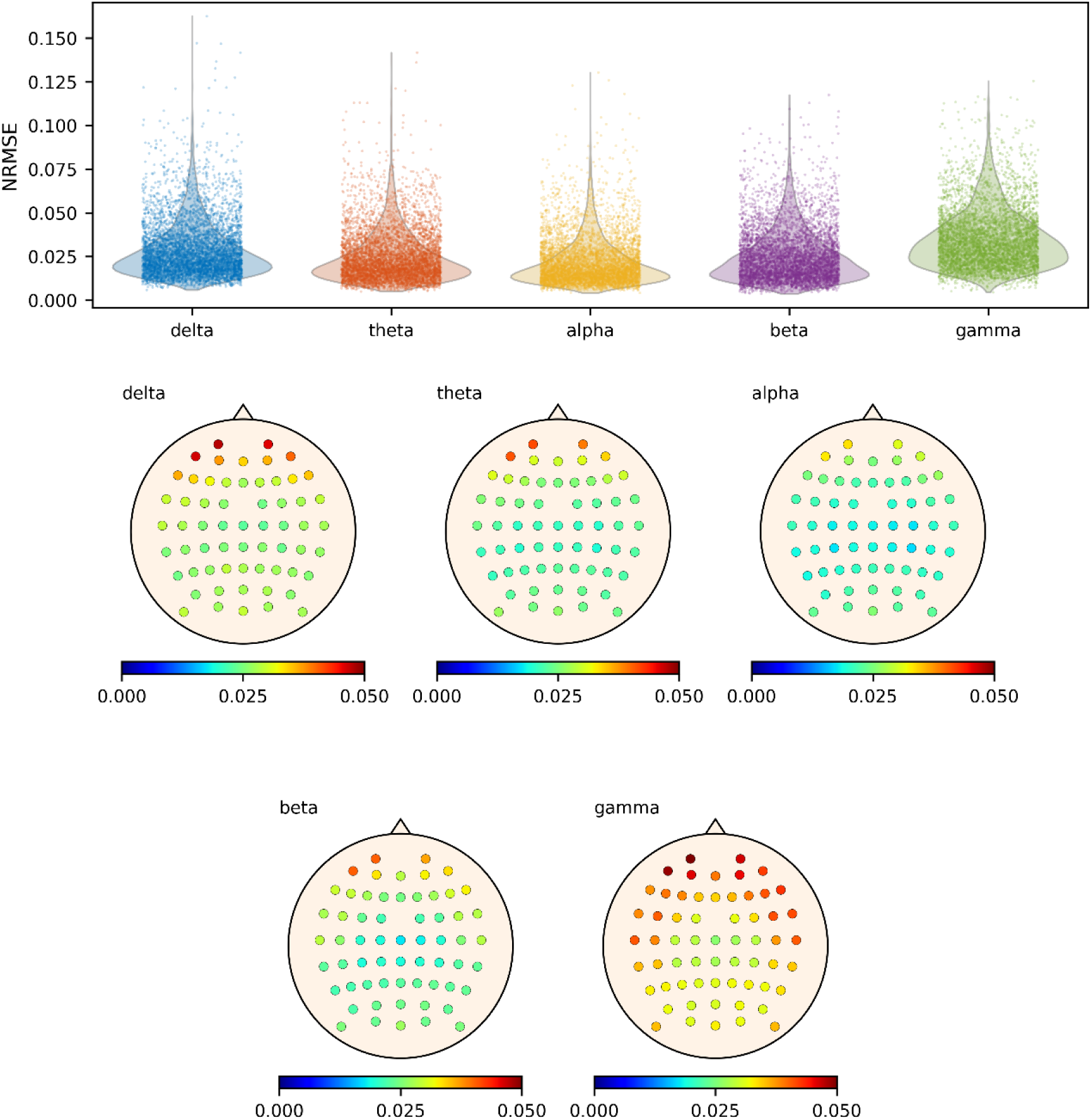
NRMSE values of PLV in the LEMON dataset. On the top panel, violin plots representing of the NRMSE distributions for all channels for each frequency band. On the bottom panels, spatial distribution of NRMSE values across the head for each frequency band. Abbreviations: NRMSE, normalised root mean square error.

#### 4.3.2. AI-Mind dataset

The PLV obtained by sEEGnal showed a great resemblance to the ones averaged from the EEG experts (Figure 21). The global resemblance is presented as the PLV of each frequency band obtained by sEEGnal for all subjects and channels against the ones obtained by the EEG experts. The correlation between sEEGnal and human experts was significant for all bands (p < 0.0001) with the lowest correlation value observed in delta band (rho = 0.8934).

**Figure 21.**
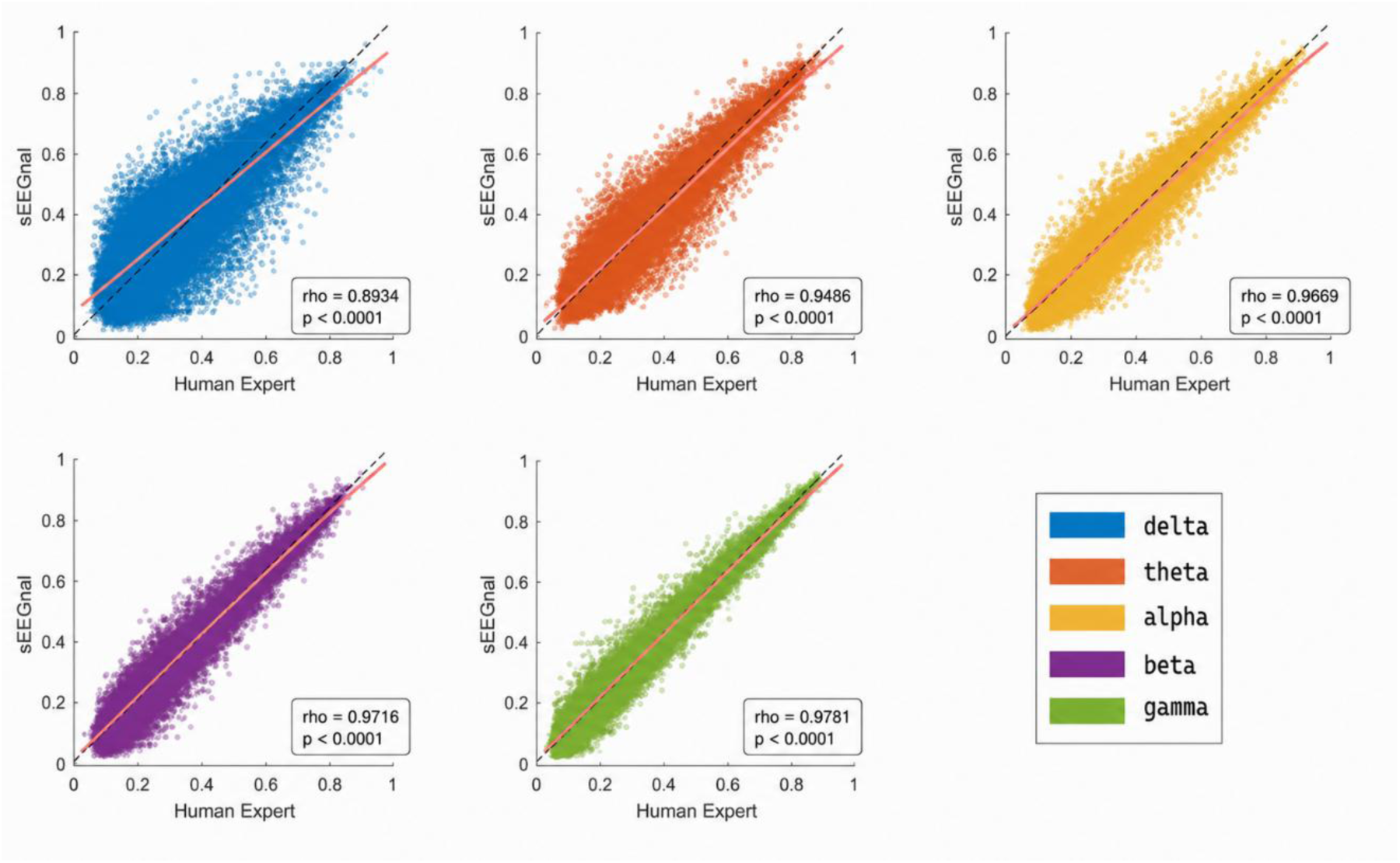
Comparison of PLV values obtained by the EEG experts and by sEEGnal in the AI-Mind dataset. An X-Y plot of the PLV values obtained in each band. The dash line represents a perfect correspondence (1:1), and the pink line represents the least-squares line. Abbreviations: PLV, phase locking value.

Regarding the frequency bands, the NRMSE values for all channels in all recordings analysed are below 0.12, meaning the maximum difference across all the data analysed is below 12 % of the metric range, with the delta band being the one with largest differences. (Figure 22, top). Regarding the spatial distribution of the differences, all channels present NRMSE values below 3.8 % of the range of the metric (Figure 22, bottom panel), with the frontal channels presenting the highest differences, especially in the delta band.

**Figure 22.**
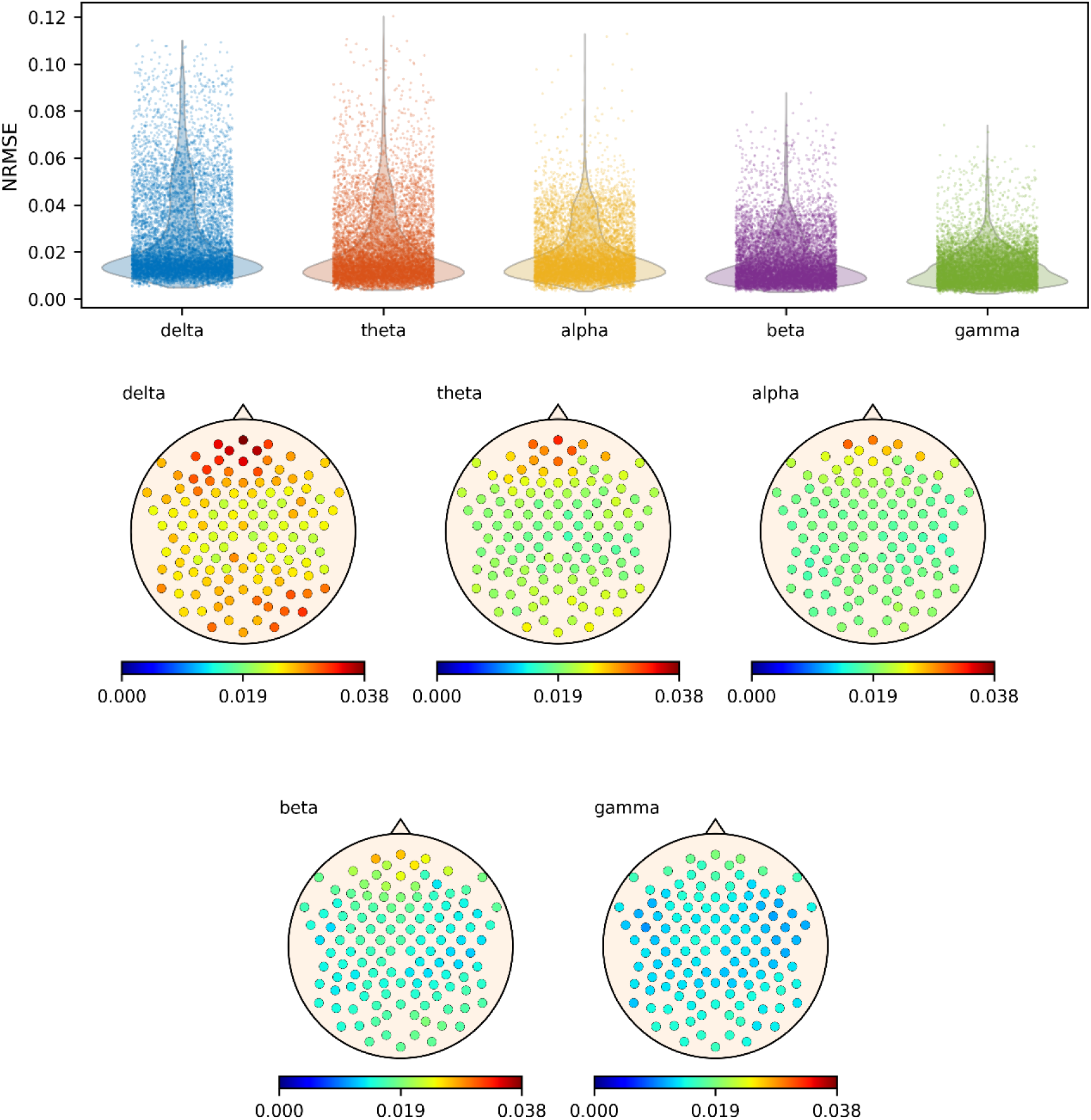
NRMSE values of PLV in the AI-Mind dataset. On the top panel, violin plots representing of the NRMSE distributions for all channels for each frequency band. On the bottom panels, spatial distribution of NRMSE values across the head for each frequency band. Abbreviations: NRMSE, normalised root mean square error.

#### 4.3.3. Test-retest dataset

For the sake of clarity, only the results for the eyes-closed task-free data are presented in this section, with those for eyes-open presented in the Supplementary Material.

The comparison of the PLV values obtained from the first and second recording of the same session are presented in Figure 23. The global resemblance is presented as the PLV of each frequency band obtained by sEEGnal for all subjects and channels comparing the first recording and second recording on the same session in an X-Y plot. The correlation between the first and second recording was significant for all bands (p < 0.0001) with the lowest correlation value observed in delta band (rho = 0.9613).

**Figure 23.**
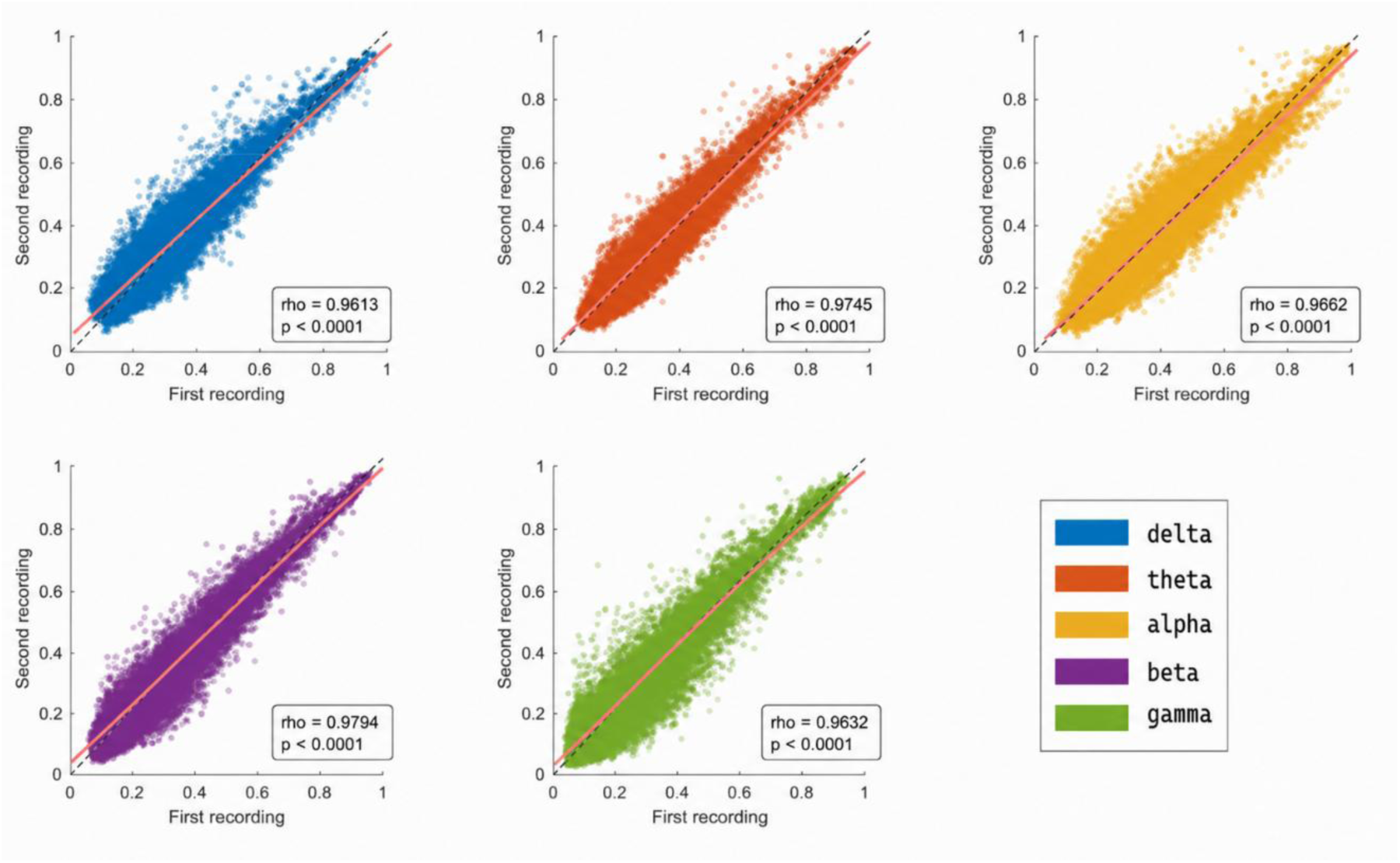
Comparison of PLV values obtained by sEEGnal on two recordings on the same session. An X-Y plot of the values obtained by sEEGnal for both recordings. The dash line represents a perfect correspondence (1:1), and the pink line represents the least-squares line.

Regarding the results per frequency band, the NRMSE values for all channels in all recordings analysed are below 0.175, meaning the maximum difference across all the data analysed is below 17.5 % of the metric range, with the alpha band being the one with largest differences. (Figure 24, top). Regarding the spatial distribution of the differences, all channels present NRMSE values below 3.4 % of the range of the signal (Figure 24, bottom panel), with the frontal channels presenting the largest differences in the alpha band, followed by frontal and occipital channels presenting major differences in the gamma band.

**Figure 24.**
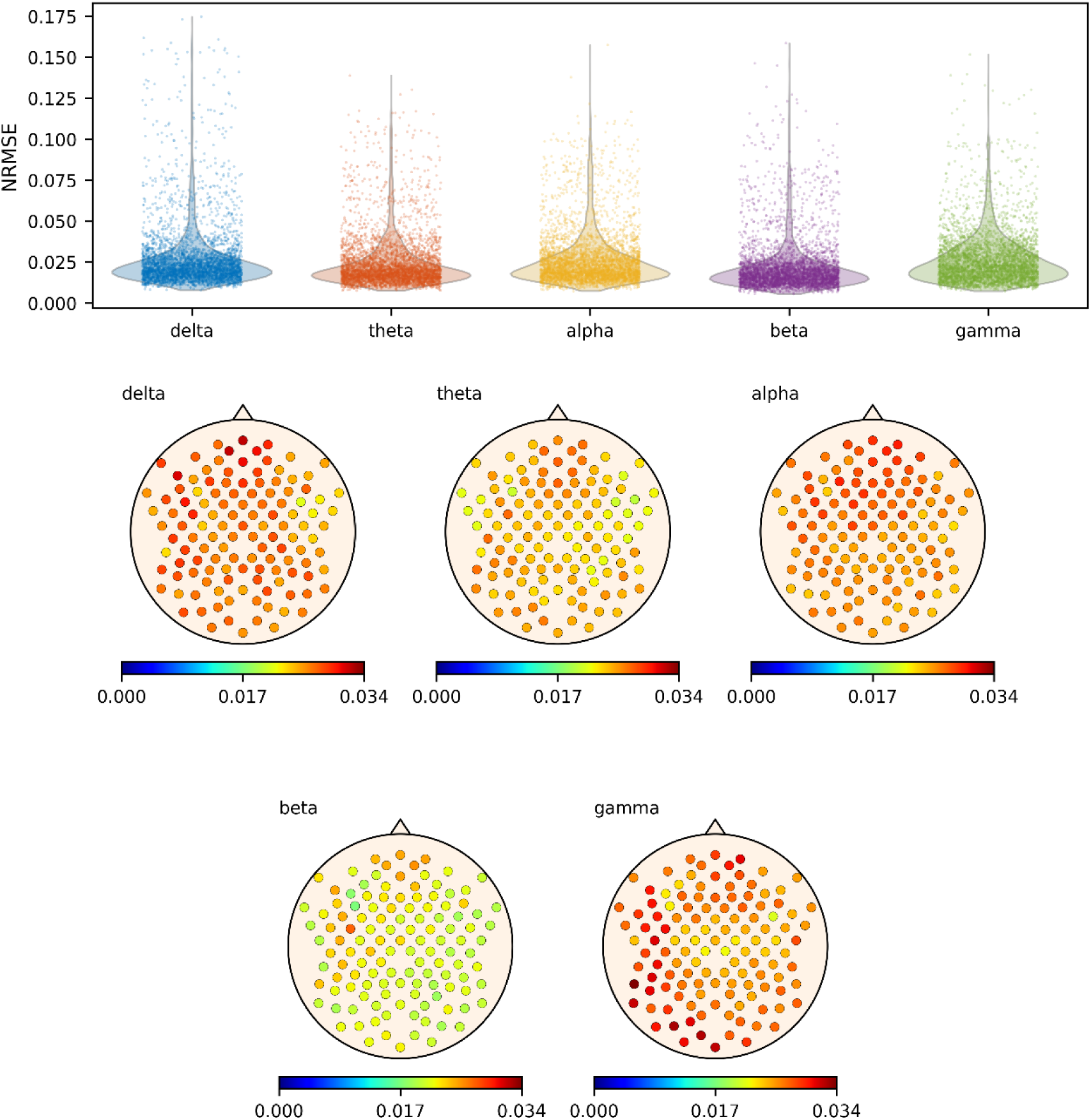
NRMSE values of PLV in the test-retest dataset. On the top panel, violin plots representing of the NRMSE distributions for all channels for each frequency band. On the bottom panels, spatial distribution of NRMSE values across the head for each frequency band. Abbreviations: NRMSE, normalised root mean square error.

## 5. Discussion

In this work, we present sEEGnal, an automatic and comprehensive pipeline for EEG pre-processing. The pipeline is designed as a modular and scalable framework for automatic EEG preprocessing. sEEGnal comprises three high-level modules (standardisation, bad channel detection, and artefact detection) that follow widely accepted guidelines for EEG pre-processing. sEEGnal has been designed oriented to five vital characteristics for this type of algorithm: i) being a comprehensive EEG pre-processing pipeline; ii) being fully automatic, i.e. users without programming background can use it; iii) being, from a performance point of view, comparable to expert-driven preprocessing; iv) being time-and memory-efficient for working in large datasets; and v) being open-source. Importantly, the pipeline is evaluated against expert-driven preprocessing and in terms of preservation of downstream neurophysiological measures.

The first module, standardisation, was designed following the BIDS standard[25]. This decision aligns with two of the abovementioned characteristics (comprehensiveness and open source) since BIDS is nowadays the current standard protocol to share large EEG datasets. Moreover, this design facilitates interoperability and reproducibility across multi-centre studies and heterogeneous datasets. The bad channel and artefact detection modules rely on the classification of independent components by means of ICLabel[11]. The bad channel criteria assess the key features of bad channels, namely high impedance, physiologically implausible amplitudes (both high and low amplitudes), abnormal power spectrum, gel bridges, and amplitudes deviating from amplitude variance ranges. The artefacts are classified into the common types of artefacts, namely eye-related, muscle-related, sensor-related, and “other”. Since artefact detection is also based on IC classification, the pipeline was specifically designed to detect muscle-and sensor-related artefacts first, which could otherwise dominate the ICs and result in the ocular and “other” artefacts being missed.

For this work, we aimed to evaluate the performance of sEEGnal, benchmarked against the performance of EEG experts when pre-processing real EEG recordings. To this end, we evaluated sEEGnal performance based on the bad channel and artefact detection metadata, neurophysiological measurements commonly used in EEG experiments, and the time and memory required to pre-process the recordings. This multi-level evaluation framework allows performance to be interpreted not only in terms of preprocessing decisions but also in terms of preservation of downstream neurophysiological properties.

Regarding the metadata, the results showed that the performance of sEEGnal can be compared to that of EEG experts. The first analyses did not demonstrate significant differences in the number of bad channels, artefacts, or rejected ICs.

Further analyses regarding bad channels showed similar spatial distributions of the most common bad channels, with fronto-temporal channels predominating. In addition, the Bland–Altman and Dice overlap analyses showed that the variability between sEEGnal and EEG experts was comparable to the inter-expert variability observed between human experts.

Further analyses regarding artefacts showed differences in classification: sEEGnal identified proportionally more eye-and muscle-related artefacts, whereas EEG experts showed a higher proportion of electronic jumps and artefacts classified as “other”. The temporal Dice overlap analysis further showed that the agreement strongly depended on the artefact category, with muscle and jump artefacts presenting higher temporal overlap than EOG and “other” artefacts. Importantly, the agreement variability observed between sEEGnal, and EEG experts remained within the variability observed between human experts.

When assessing the number of rejected components, both sEEGnal and EEG experts rejected a comparable number of components. Bland–Altman analyses also revealed minimal systematic bias and agreement patterns comparable to the inter-expert variability.

A point worth discussing is the potential circularity associated with the use of ICLabel, which was originally trained using human-labelled ICs. Therefore, the observed results should be interpreted both as good performance on ICLabel part and as consistency with expert-driven component rejection rather than as a completely independent validation of IC classification.Nevertheless, it is important to note that the EEG experts involved in the present study were fully independent from the development and training of ICLabel and had never previously used the tool, and followed a completely different criteria to label components, avoiding which disregards any overlap between training and validation annotations. In addition, the evaluation framework extended beyond IC-level agreement, aits included bad channel and artefact metadata, power spectral density, functional connectivity, and test–retest consistency.

Altogether, these results suggest that sEEGnal successfully captures the main characteristics of bad-quality EEG data while preserving relevant neurophysiological properties prior to downstream analyses.

To draw better conclusions, not only the metadata should be analysed. In this study, we estimated and analysed two common features used in neurophysiological studies, namely power spectrum and functional connectivity (by means of PLV)[26]. This approach allowed us to assess whether sEEGnal is valid for use in neuroscientific works. Regarding the power analysis, the power spectra obtained by sEEGnal resembles with high accuracy those obtained by EEG experts both in the AI-Mind and LEMON datasets. The largest difference was found in the gamma band, which is a band with a low signal-to-noise ratio, and might be affected by differences in the level of detection of high-frequency muscle-related artefacts. When analysing the spatial distribution of the differences, i.e. analysing the performance of each channel, we found an overall good agreement, finding the largest differences over the frontal channels in the delta band. Since both eyes-open and eyes-closed task-free recordings were analysed together, this difference may be related to differences in the identification of eye-related artefacts and/or components. Nevertheless, we observed that the differences’ magnitude is low compared to the range of the feature (around 8 %).

For functional connectivity, we found overall good agreement, but with higher deviations than in the power spectrum analysis. This was expected when considering the nature of both neurophysiological measures; the power spectrum being one of the most robust measures (and so it is reflected in the results), and functional connectivity far more dependent on the pre-processing decisions[27,28]. Regarding the resemblance, i.e. comparing the absolute PLV values, the PLV values obtained after applying sEEGnal are consistently lower in all frequency bands, especially in the delta and gamma band. When analysing the results by sensors, frontal channels were the ones with largest differences. Again, this difference may be related to eye-related artefacts and/or components disagreement between sEEGnal and experts, which would affect PLV to a greater extent than spectral power.

Another key feature to analyse is the consistency of the neurophysiological metrics obtained after applying sEEGnal[29–31]. For that end, we selected different recordings from the same session of each subject. It was expected that recordings from the same subject recorded within a 10-minute period would present similar neurophysiological outcomes. For power analysis, the results showed an overall good consistency, with the largest differences in frontal channels in delta band for open-eyes and in occipital channels in alpha band for closed-eyes. These differences were expected due to the nature of power spectrum, with eye movements affecting the delta band[32] and with very large differences between eyes-open and eyes-closed conditions in the alpha band[33].

Regarding the time required to pre-process a single EEG recording, there were no significant differences in the average time required to complete each module between sEEGnal and EEG experts. In contrast, when estimating the standard deviation of the time used for a single EEG recording, the EEG experts presented a much higher variability, indicating poorer consistency than sEEGnal. In addition, sEEGnal did not present problems associated with humans, like tiredness or available working hours. The results and the nature of the algorithm make sEEGnal a robust solution for pre-processing large EEG datasets.

Importantly, this open-source algorithm has been tested in two different datasets, one public and one private, using devices from two different manufacturers and different configurations (such as the number of channels, the sample frequency, or the length of the recordings). These results highlight the potential of sEEGnal to generalize across different hardware setups and acquisition protocols.

In recent years, several automated EEG preprocessing pipelines have been proposed, each prioritizing different aspects of the preprocessing workflow. For example, FASTER [34] focuses on fully automated statistical detection of outlier channels, epochs, and components; Autoreject [35] emphasizes adaptive threshold estimation and automated repair strategies; HAPPE [36] was specifically designed for highly contaminated recordings and developmental populations; RELAX [37] combines multiple automated cleaning approaches to optimize artefact removal; and DISCOVER-EEG [38] provides a standardised and reproducible preprocessing workflow for large-scale EEG studies. Although these pipelines substantially reduce manual intervention, many still require users to interact with MATLAB, EEGLAB, FieldTrip, MNE-Python, or command-line workflows. Rather than competing in terms of maximizing artefact attenuation or outperforming existing solutions, sEEGnal was designed to provide a transparent, modular, and reproducible preprocessing framework while minimising programming requirements through the use of configuration files rather than user-written code. In particular, the pipeline integrates BIDS-based standardisation, physiologically grounded criteria for bad channel and artefact detection, and validation relative to expert-driven preprocessing and downstream neurophysiological measures. This design aims to facilitate scalable, interpretable, and user-accessible EEG preprocessing across heterogeneous datasets and experimental settings.

Although the results in this work demonstrate that the outcomes of sEEGnal are overall comparable to those of human expert pipelines, some limitations merit discussion. Firstly, for this work we had access to only two datasets, limiting the validation of sEEGnal in a wider variety of EEG manufacturers. Nevertheless, the two datasets used in this study have sufficient quality to support the validity of sEEGnal. The same applies to the reduced number of EEG experts. Secondly, the current version of sEEGnal heavily relies on the use of external packages, especially ICLabel[11], with the intrinsic risk of depending on external packages. Nevertheless, ICLabel is a widely used tool, and uses machine learning algorithms trained on expert-labelled ICs derived from large EEG datasets. It would be unfeasible, and extremely inefficient, to try and replicate this effort independently for sEEGnal. Finally, sEEGnal has been only evaluated in resting-state data. An evaluation of sEEGnal in task-related EEG experiment would strengthen the proof-of-concept of sEEGnal.

In conclusion, sEEGnal represents a robust, fully automatic, and comprehensive pipeline for EEG pre-processing that achieves performance comparable to that of expert EEG practitioners. Its modular design, adherence to widely accepted standards, and ability to manage diverse datasets efficiently make it a versatile tool for both small-and large-scale studies. The pipeline reliably identifies bad channels, artefacts, and independent components, while preserving the integrity of key neurophysiological measures such as spectral power and functional connectivity. Moreover, sEEGnal demonstrates superior consistency and reproducibility compared to human experts, and its open-source nature ensures accessibility and adaptability for the broader neuroscience community.

Collectively, these features establish sEEGnal as a robust preprocessing framework for automatic EEG pre-processing.

## 6. Code availability

The updated source code of sEEGnal may be found in https://github.com/FedeC3N/sEEGnal.

The sEEGnal version and all the scripts developed in this study for testing sEEGnal may be found in https://github.com/FedeC3N/sEEGnal_perfomance. sEEGnal is coded mainly in Python (with two modules coded in C).

The statistical scripts and the visualization scripts were coded in MATLAB by the authors of this paper.

All external packages and their versions can be found on Supplementary Material.

The config file with all the parameters used in this work can be found in Supplementary Material.

## Data Availability

The updated source code of sEEGnal may be found in https://github.com/FedeC3N/sEEGnal.
The sEEGnal version and all the scripts developed in this study for testing sEEGnal may be found in https://github.com/FedeC3N/sEEGnal_perfomance.

## 8. Acknowledgment

The AI-Mind dataset is stored on the *Tjenester for Sensitive Data* (TSD) facilities, owned by the University of Oslo, operated and developed by the TSD service group at the University of Oslo, IT-Department (USIT).

## 9. Authors contribution

Federico Ramírez-Toraño: conceptualization, data curation, formal analysis, writing – original draft, writing – review and editing. Christoffer Hatlestad-Hall: conceptualization, data curation, writing – review and editing. Ainar Drews: project administration, resources. Hanna Renvall: conceptualization, writing – review and editing. Paolo María Rossini: conceptualization, writing – review and editing. Camillo Marra: conceptualization, writing – review and editing. Ira H. Haraldsen: conceptualization, writing – review and editing. Fernando Maestu: conceptualization, writing – review and editing. Ricardo Bruña: conceptualization, data curation, supervision, writing – original draft, writing – review and editing.

## 10. Ethics statement

This study is based on secondary analysis of previously collected data from human participants. All original data acquisition procedures were conducted in accordance with the Declaration of Helsinki and complied with relevant national and institutional guidelines. Ethical approval was obtained by the corresponding local ethics committees at each participating site within the framework of the study protocols (including approval dates and reference numbers as specified in the original studies).

All participants provided written informed consent prior to participation, including consent for the use of their data in secondary analyses. The present work did not involve any new data collection.

All data used in this study were fully anonymized prior to analysis, and no identifiable personal information was accessed. The privacy and confidentiality of participants were strictly maintained throughout all stages of the study.

