## Supplementary Material for "sEEGnal: an automated EEG preprocessing pipeline evaluated against expert-driven preprocessing"

### Supplementary Materials

#### Impedances distribution

There is no clear consensus regarding acceptable impedance values for EEG channels. Previous studies have shown that higher electrode impedance does not directly determine signal quality[1]. In this study, we defined a heuristic impedance threshold and subsequently evaluated its validity against expert annotations.

We estimated and visualized the distribution of impedance values across recordings. We then examined the relationship between channel impedance and the likelihood of being marked as bad by EEG experts. No association was observed between impedance and bad channel classification below the selected threshold. In contrast, the four channels with impedance values above the threshold were consistently identified as bad by all experts.

Preprint

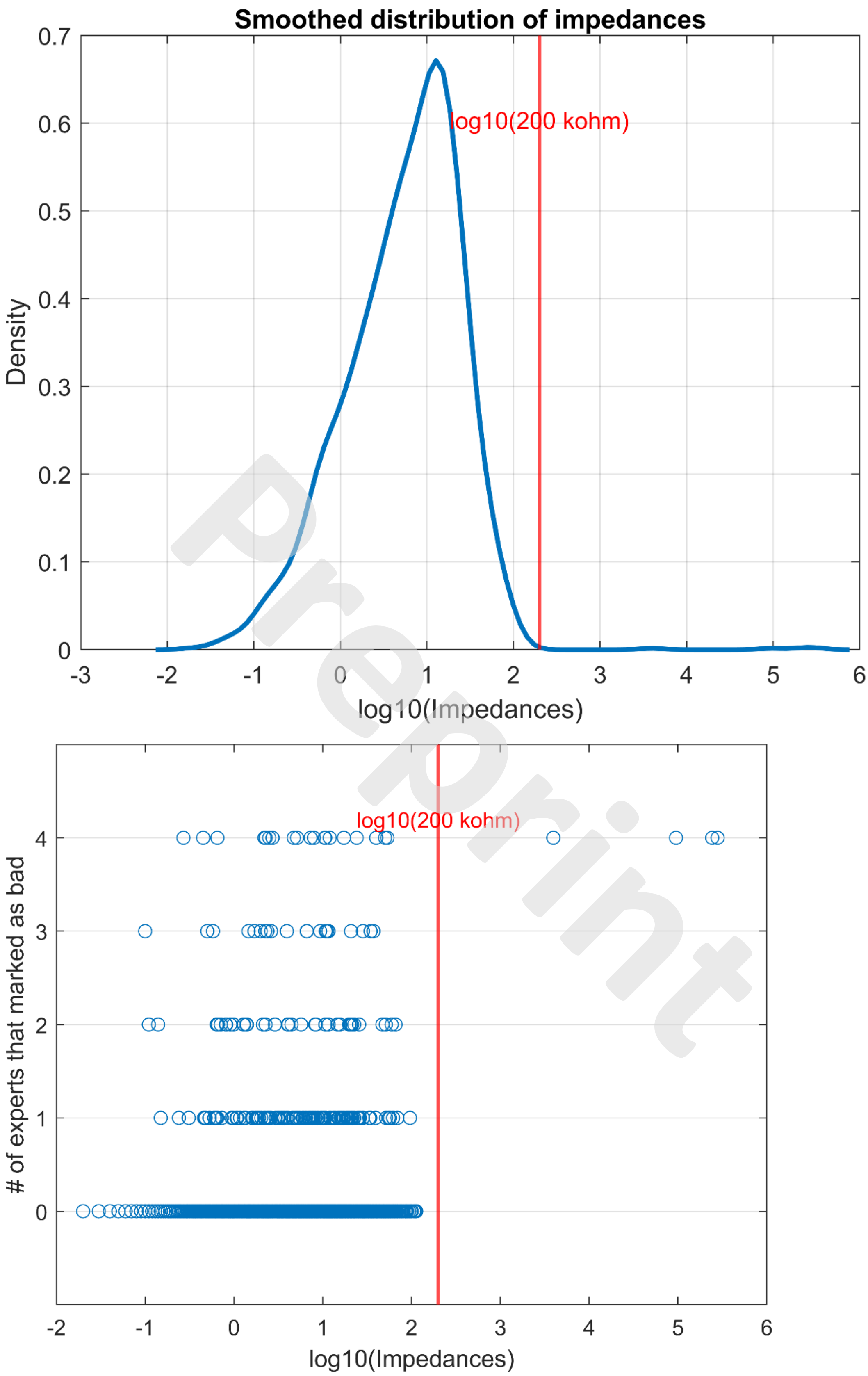


Preprint

Figure S1. Distribution of impedances. On the top panel, the distribution of impedances for all channels in the 20 EEG recordings.

On the bottom panel, for each channel, the number of expert that marked the channel as bad and its impedance. The red line indicates the threshold of 200 kΩ in logarithmic scale.

#### Power spectrum criteria

During the development of sEEGnal, multiple criteria and thresholds were explored. Some of the final thresholds were selected based on heuristic considerations. In the pipeline used by EEG experts, signals are visually inspected after band-pass filtering between 2 and 45 Hz. We observed that some channels classified as bad exhibited increased power at higher frequencies outside the visualization range.

Figure S2 presents the channels identified by sEEGnal using this criterion and the number of EEG experts who also classified those channels as bad. Although some false positives were observed (crosses on the bottom line represent channels marked as bad by sEEGnal but not by any expert), most of the channels identified by this criterion were also marked as bad by the EEG experts.

Preprint

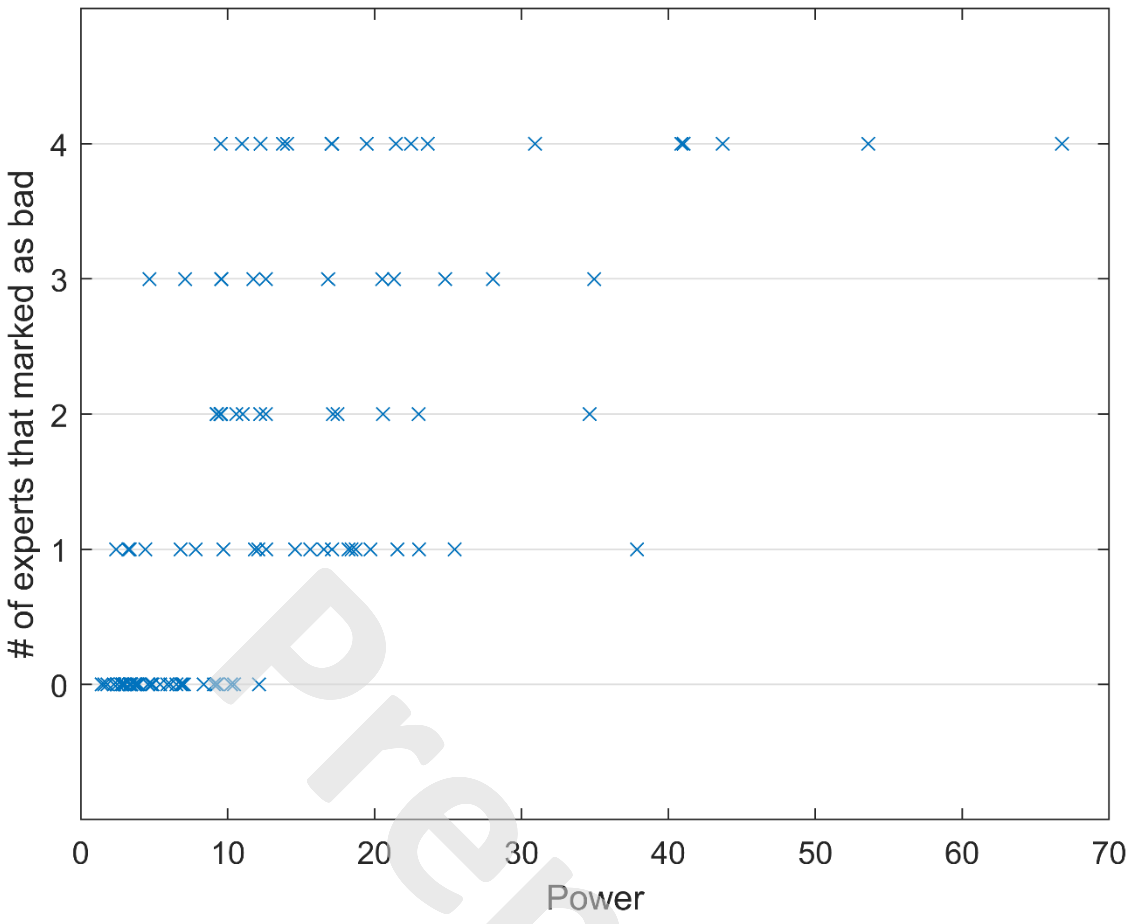


Preprint

#### BIDS structure

BIDS standard describes a way of arranging data and specifying metadata for a subset of neuroimaging experiments. It follows a simple but carefully defined terminology. The filenames are formed with a series of key-values and end with a file type, where keys and file types are predefined, and values are chosen by the user.

BIDS standard aims to: ease the data sharing; reduce the number of errors due to the misunderstanding of the meaning of a given datum; optimize the usage of data analysis software. Figure S1 presents the folder structure used in this project.

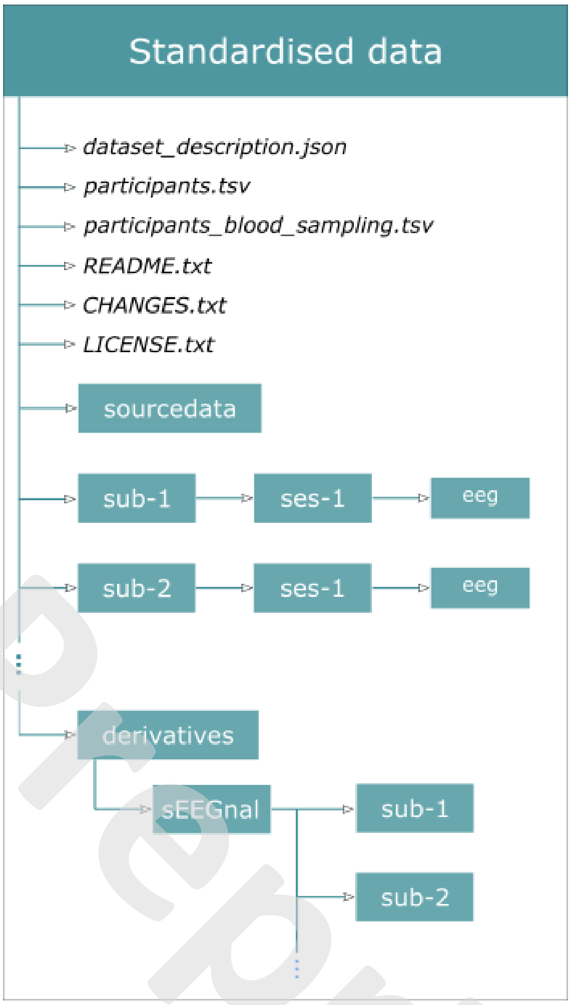


Figure S3. Folder structure in AI-Mind following the BIDS protocol.

Preprint

#### sEEGnal package requirements

As defined in requirements.txt:

- aimind.meeg>=0.4.2
- eeglabio>=0.0.3
- mne>=1.9.0
- mne-bids>=0.12.0
- mne-icalabel>=0.4
- numpy>=1.23
- scipy

- pandas
- pybv>= 0.7.3
- torch

Preprint

#### sEEGnal config parameters

The preprocessing parameters of sEEGnal are defined through editable JSON configuration files. This design allows users to modify thresholds, filtering ranges, epoch definitions, and artifact-detection criteria without editing the source code. The following configuration corresponds to the default parameters used in the present study.

| Module | Submodule | Parameter | Default value | Description |
|---|---|---|---|---|
| Visualization | General | low_freq_limit | 2 Hz | Lower frequency limit for visualization |
| | | high_freq_limit | 45 Hz | Upper frequency limit for visualization |

| Module | Submodule | Parameter | Default value | Description |
|---|---|---|---|---|
| Component estimation | Preprocessing | low_freq | 1 Hz | Lower bandpass filter cutoff before ICA |
| | | high_freq | 100 Hz | Upper bandpass filter cutoff before ICA |
| | | resampled_frequency | 500 Hz | Resampling frequency before ICA |
| | | notch_frequencies | 50–250 Hz | Frequencies removed with notch filters |
| | ICLabel | unclear_threshold | 0.7 | ICLabel uncertainty threshold |
| | Epoching | length | 4 s | Epoch duration |
| | | overlap | 0 s | Epoch overlap |
| | | padding | 2 s | Padding added to each epoch |

| Module | Submodule | Parameter | Default value | Description |
|---|---|---|---|---|
| Bad channel detection | General | crop_seconds | 10 s | Initial crop to reduce edge effects |
| | High impedance | threshold | 200 kΩ | Impedance threshold |
| | Impossible amplitude | low_freq | 2 Hz | Lower frequency bound |
| | | high_freq | 150 Hz | Upper frequency bound |
| | | low_threshold | 1 µV | Minimum acceptable amplitude |

Preprint

| | | | | |
|---|---|---|---|---|
| | | high_threshold | 500 µV | Maximum acceptable amplitude |
| | | percentage_threshold | 0.5 | Required proportion of affected epochs |
| | Power spectrum | low_freq | 45 Hz | Lower frequency bound |
| | | high_freq | 55 Hz | Upper frequency bound |
| | | threshold | 3 | Spectral power ratio threshold |
| | | percentage_threshold | 0.5 | Required proportion of affected epochs |
| | Gel bridge | low_freq | 8 Hz | Lower frequency bound |
| | | high_freq | 45 Hz | Upper frequency bound |
| | | threshold | 0.999 | Correlation threshold |
| | | neighbour_distance | 0.05 m | Maximum distance between neighbouring electrodes |
| | | seq_threshold | 0.5 | Required proportion of affected epochs |
| | High deviation | low_freq | 2 Hz | Lower frequency bound |
| | | high_freq | 45 Hz | Upper frequency bound |
| | | threshold | 3 | Variance ratio threshold |
| | | percentage_threshold | 0.5 | Required proportion of affected epochs |

| Module | Submodule | Parameter | Default value | Description |
|---|---|---|---|---|
| Artifact detection | Muscle | low_freq | 110 Hz | Lower frequency bound |
| | | high_freq | 145 Hz | Upper frequency bound |

Preprint

| | | | | |
|---|---|---|---|---|
| | | threshold | 10 | Peak amplitude ratio threshold |
| | | crop_seconds | 10 s | Initial crop |
| | | resampled_frequency | 500 Hz | Resampling frequency |
| | Sensor | low_freq | 0.5 Hz | Lower frequency bound |
| | | high_freq | 2 Hz | Upper frequency bound |
| | | threshold | 0.99 | Correlation threshold |
| | | ratio | 5 | Peak-to-standard-deviation ratio |
| | | crop_seconds | 10 s | Initial crop |
| | | resampled_frequency | 500 Hz | Resampling frequency |
| | Other | low_freq | 2 Hz | Lower frequency bound |
| | | high_freq | 45 Hz | Upper frequency bound |
| | | threshold | 500 µV | Maximum acceptable amplitude |
| | | crop_seconds | 10 s | Initial crop |
| | | resampled_frequency | 500 Hz | Resampling frequency |
| | EOG | low_freq | 2 Hz | Lower frequency bound |
| | | high_freq | 5 Hz | Upper frequency bound |
| | | ratio | 10 | Frontal/background activity ratio |
| | | crop_seconds | 10 s | Initial crop |
| | | resampled_frequency | 500 Hz | Resampling frequency |

| Module | Submodule | Parameter | Default value | Description |
|---|---|---|---|---|
| Export clean EEG | General | crop_seconds | 10 s | Initial crop |
| | | notch_frequencies | 50–250 Hz | Frequencies removed with notch filters |
| | Epoching | length | 4 s | Epoch duration |
| | | overlap | 0 s | Epoch overlap |
| | | padding | 2 s | Padding added to each epoch |

Preprint

#### Pseudocode of statistical analysis

For the sake of clarity, a pseudocode of the methodology used in the statistical analysis of power spectrum and PLV is presented in Figure S4.

```
# First, load all the measures for sEEGnal and the human EEG expert
load sEEGnal measure;
load EEG expert measure;

for each frequency band:

    select frequencies in sEEGnal measure;
    select frequencies in EEG expert measure;

    for each sensor:

        select sensor in sEEGnal measure;
        select sensor in EEG expert measure;

        for each recording:

            # Estimate correlation, and NRMSE
            corr(sEEgnal measure(freq,sensor,subject),EEG expert measure(freq,sensor,subject))
            NRMSE(sEEgnal measure(freq,sensor,subject),EEG expert measure(freq,sensor,subject))

        # Average across recordings for each sensor
        mean(corr)
        mean(NRMSE)

    # Plot the spatial results for each frequency band
    plot_head(corr)
    plot_head(NRMSE)
```

Figure S4. Pseudocode of statistical analysis.

Preprint

#### Bland-Altman analysis

Bland–Altman agreement analyses for all pairwise preprocessing comparisons, including sEEGnal versus each individual EEG expert and expert-versus-expert comparisons. These analyses were performed for the number of bad channels, artefacts, and rejected ICs. The supplementary figures allow the visualization of the variability associated with each individual comparison and provide a detailed characterization of the agreement patterns underlying the summarized results presented in the main manuscript.

Preprint

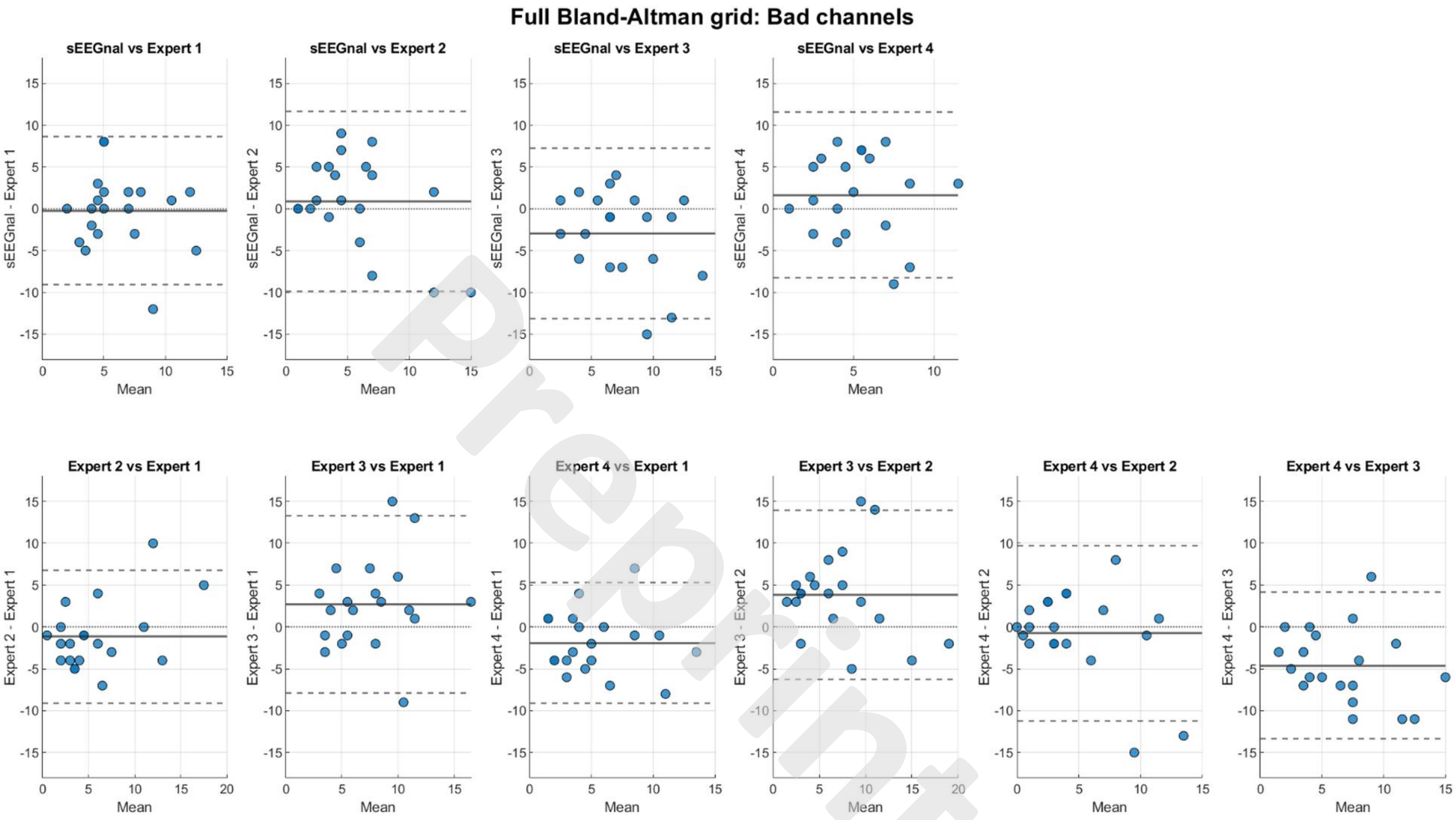
Full Bland-Altman grid: Bad channels
sEEGnal vs Expert 1
sEEGnal vs Expert 2
sEEGnal vs Expert 3
sEEGnal vs Expert 4
sEEGnal - Expert 1
sEEGnal - Expert 2
sEEGnal - Expert 3
sEEGnal - Expert 4
Expert 2 vs Expert 1
Expert 3 vs Expert 1
Expert 4 vs Expert 1
Expert 3 vs Expert 2
Expert 4 vs Expert 2
Expert 4 vs Expert 3
Expert 2 - Expert 1
Expert 3 - Expert 1
Expert 4 - Expert 1
Expert 3 - Expert 2
Expert 4 - Expert 2
Expert 4 - Expert 3
Mean

Preprint

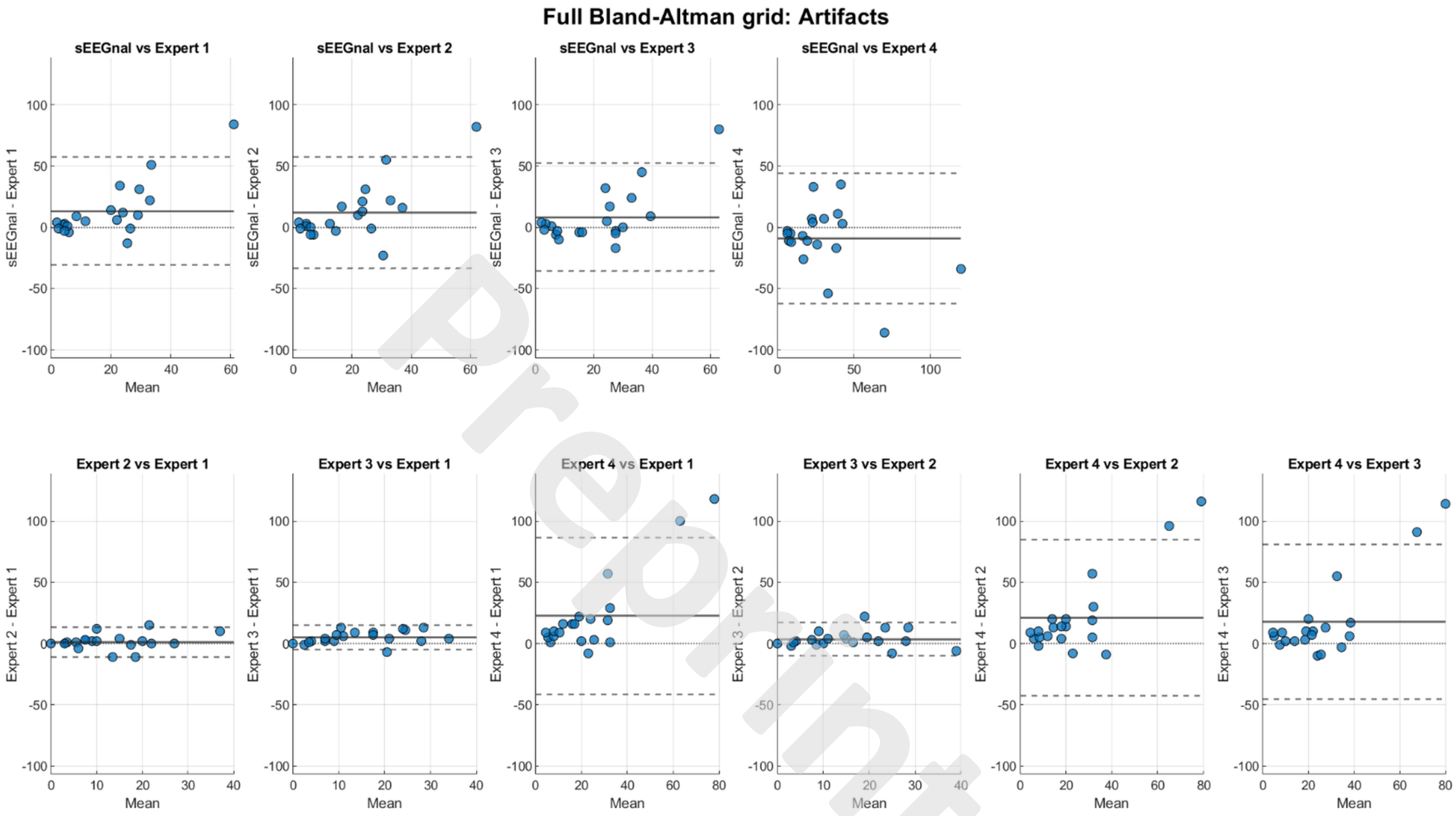
Full Bland-Altman grid: Artifacts
sEEGnal vs Expert 1
sEEGnal vs Expert 2
sEEGnal vs Expert 3
sEEGnal vs Expert 4
sEEGnal - Expert 1
sEEGnal - Expert 2
sEEGnal - Expert 3
sEEGnal - Expert 4
Expert 2 vs Expert 1
Expert 3 vs Expert 1
Expert 4 vs Expert 1
Expert 3 vs Expert 2
Expert 4 vs Expert 2
Expert 4 vs Expert 3
Expert 2 - Expert 1
Expert 3 - Expert 1
Expert 4 - Expert 1
Expert 3 - Expert 2
Expert 4 - Expert 2
Expert 4 - Expert 3
Mean

Preprint

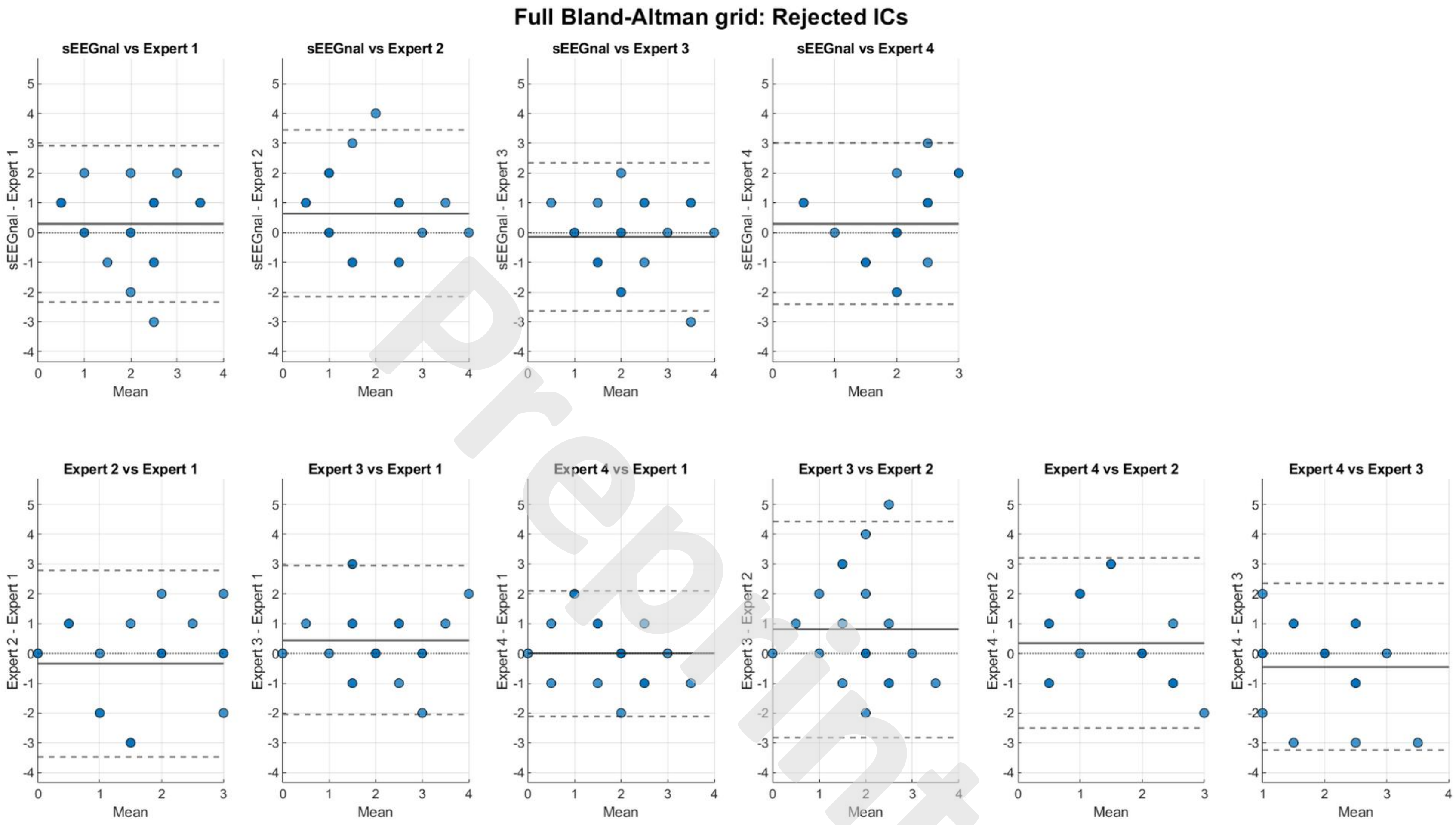
Full Bland-Altman grid: Rejected ICs
sEEGnal vs Expert 1
sEEGnal vs Expert 2
sEEGnal vs Expert 3
sEEGnal vs Expert 4
Expert 2 vs Expert 1
Expert 3 vs Expert 1
Expert 4 vs Expert 1
Expert 3 vs Expert 2
Expert 4 vs Expert 2
Expert 4 vs Expert 3
sEEGnal - Expert 1
sEEGnal - Expert 2
sEEGnal - Expert 3
sEEGnal - Expert 4
Expert 2 - Expert 1
Expert 3 - Expert 1
Expert 4 - Expert 1
Expert 3 - Expert 2
Expert 4 - Expert 2
Expert 4 - Expert 3
Mean
Preprint

#### Test-retest results

##### Power

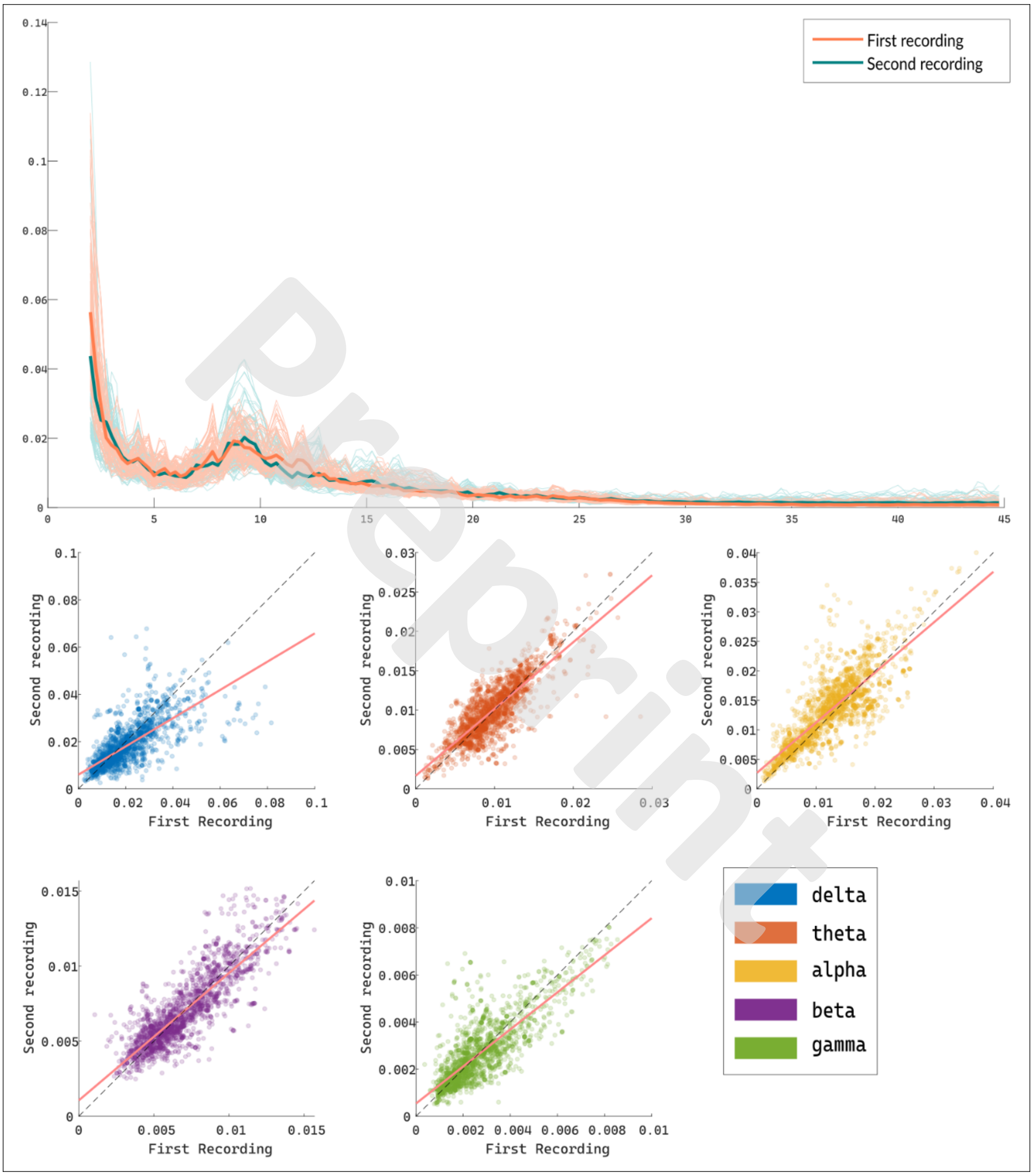


Preprint

Figure S5. Comparison of power spectra obtained by sEEGnal on the test-retest dataset. On the top panel, the power spectra of both eyes-open recordings. The pale lines represent individual channels, and the solid lines represent the average power spectrum across channels. On the bottom panel, an X-Y plot of the value obtained by sEEGnal on both recordings for the power in each band. The dash line represents a perfect correspondence (1:1), and the pink line represents the least-squares line.

Preprint

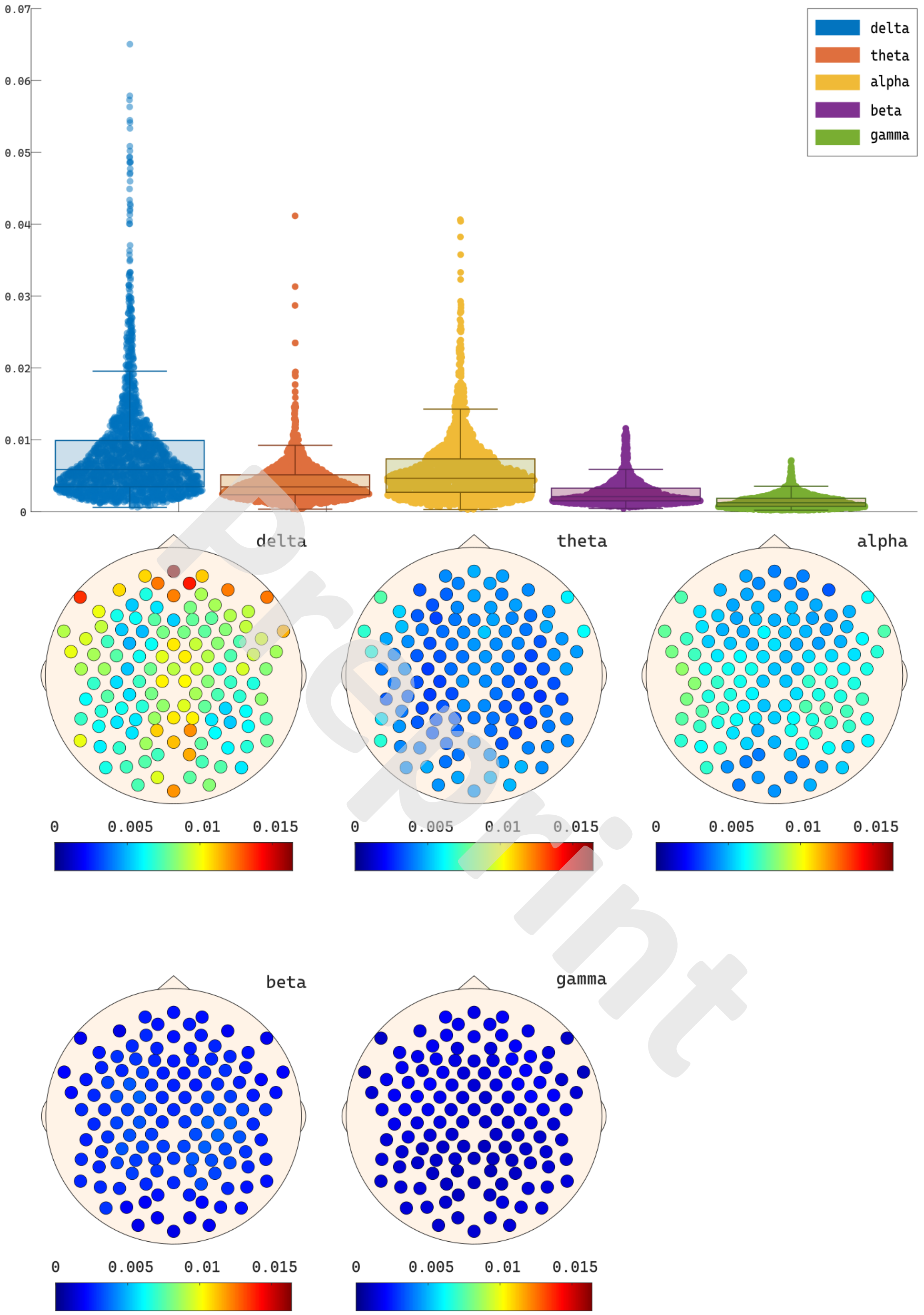


Preprint

Figure S6. NMRSE values of the power spectra in the test-retest dataset. On the top panel, violin plots representing of the NRMSE distributions for all channels for each frequency band. On the bottom panels, spatial distribution of NRME values across the head for each frequency band.

#### Functional Connectivity

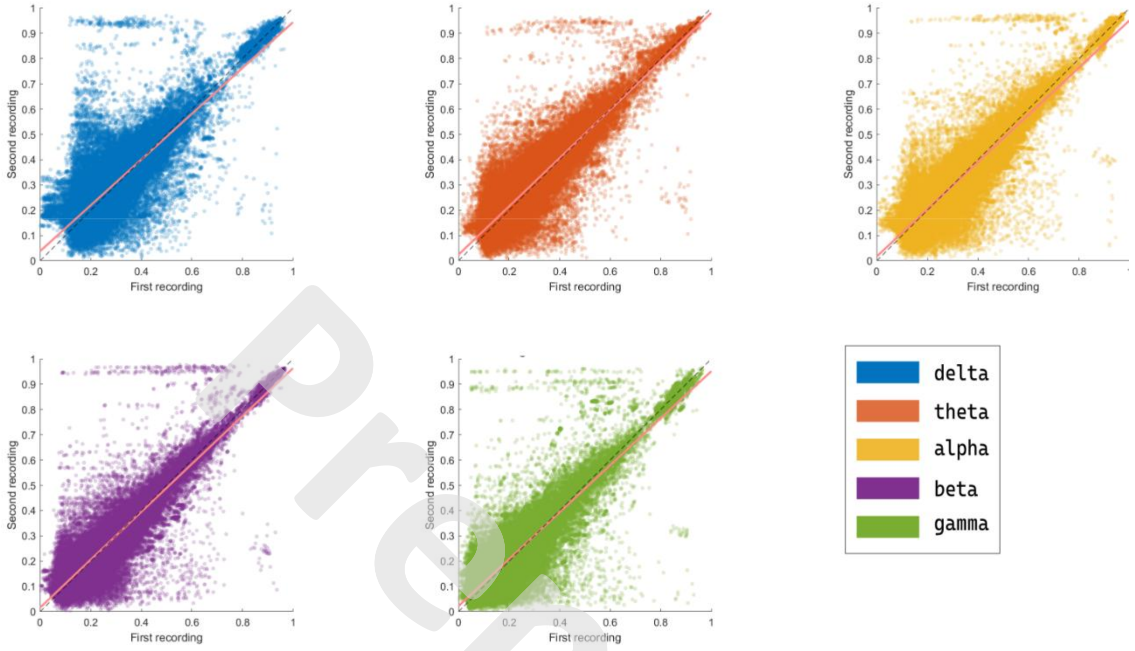


Figure S7. Comparison of PLV values obtained by sEEGnal on two eyes-open recordings on the same session. An X-Y plot of the values obtained by sEEGnal for both recordings. The dash line represents a perfect correspondence (1:1), and the pink line represents the least-squares line.

Preprint

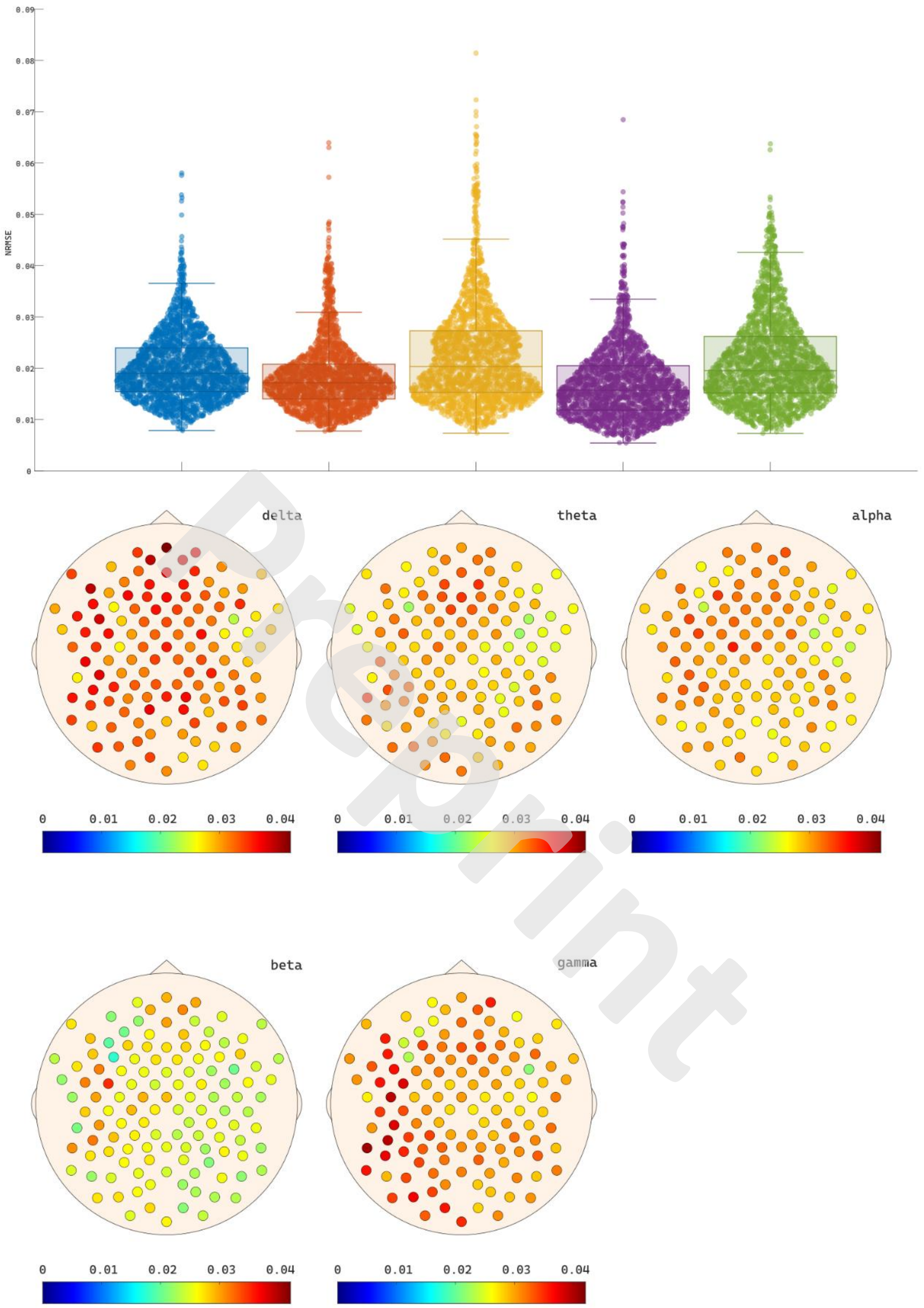


Preprint

Figure S8. NMRSE values of PLV in the test-retest dataset. On the top panel, violin plots representing of the NRMSE distributions for all channels for each frequency band. On the bottom panels, spatial distribution of NRME values across the head for each frequency band.

Preprint
